# Axonal stimulation affects the linear summation of single-point perception in three Argus II users

**DOI:** 10.1101/2023.07.21.23292908

**Authors:** Yuchen Hou, Devyani Nanduri, Jacob Granley, James D. Weiland, Michael Beyeler

## Abstract

**Purpose:** Retinal implants use electrical stimulation to elicit perceived flashes of light (“phosphenes”). Single-electrode phosphene shape has been shown to vary systematically with stimulus parameters and the retinal location of the stimulating electrode, due to incidental activation of passing nerve fiber bundles. However, this knowledge has yet to be extended to paired-electrode stimulation.

**Methods:** We retrospectively analyzed 3548 phosphene drawings made by three blind participants implanted with an Argus II Retinal Prosthesis. Phosphene shape (characterized by area, perimeter, major and minor axis length) and number of perceived phosphenes were averaged across trials and correlated with the corresponding single-electrode parameters. In addition, the number of phosphenes was correlated with stimulus amplitude and neuroanatomical parameters: electrode-retina and electrode-fovea distance as well as the electrode-electrode distance to (“between-axon”) and along axon bundles (“along-axon”). Statistical analyses were conducted using linear regression and partial correlation analysis.

**Results:** Simple regression revealed that each paired-electrode shape descriptor could be predicted by the sum of the two corresponding single-electrode shape descriptors (*p <* .001). Multiple regression revealed that paired-electrode phosphene shape was primarily predicted by stimulus amplitude and electrode-fovea distance (*p <* .05). Interestingly, the number of elicited phosphenes tended to increase with between-axon distance (*p <* .05), but not with along-axon distance, in two out of three participants.

**Conclusions:** The shape of phosphenes elicited by paired-electrode stimulation was well predicted by the shape of their corresponding single-electrode phosphenes, suggesting that two-point perception can be expressed as the linear summation of single-point perception. The notable impact of the between-axon distance on the perceived number of phosphenes provides further evidence in support of the axon map model for epiretinal stimulation. These findings contribute to the growing literature on phosphene perception and have important implications for the design of future retinal prostheses.

## 1. Introduction

Retinitis pigmentosa (RP) is an inherited degenerative disease of the eye that is estimated to affect one in 4,000 individuals worldwide (Hamel, 2006). Although recent advances in gene and stem cell therapies (e.g., Russell et al., 2017, da Cruz et al., 2018; for a recent review see McGregor, 2019) as well as retinal sheet transplants (e.g., Foik et al., 2018, Gasparini et al., 2019; for a recent commentary see Beyeler, 2019) are showing great promise as near-future treatments for early-stage RP, electronic retinal prostheses continue to be a pertinent option for later stages of the disease (Beyeler et al., 2017b).

Retinal prostheses typically acquire visual input via an external camera, which is then translated into electrical pulses sent to a microstimulator implanted in the eye (Weiland et al., 2016). The stimulator receives the information, decodes it, and stimulates the surviving retinal neurons with electrical current, thus evoking the perception of flashes of light (“phosphenes”). The most widely adopted retinal implant thus far is the Argus II Retinal Prosthesis System (Vivani Medical, Inc; formerly Second Sight Medical Products, Inc.), which was the first retinal implant to obtain regulatory approval in the US and Europe, and has been implanted in roughly 500 individuals worldwide (Luo and da Cruz, 2016).

A series of papers demonstrated that phosphenes elicited by stimulating a single Argus II electrode have a distinctive shape that is relatively consistent over time (Nanduri et al., 2008; Luo et al., 2016; Beyeler et al., 2019b). Phosphene shape has been shown to depend strongly on the retinal location of the stimulating electrode, predominantly elongated along the trajectory of the underlying nerve fiber bundle (Rizzo et al., 2003; Beyeler et al., 2019b). In addition, phosphene appearance varies systematically with stimulus amplitude and frequency (Horsager et al., 2009; Nanduri et al., 2012; Sinclair et al., 2016) to the extent that a simple computational model can predict phosphene shape across a wide range of stimulus parameters (Granley and Beyeler, 2021).

However, less is known about how phosphenes combine when multiple electrodes are stimulated. Early research suggested that repeated paired stimulation resulted in reproducible phosphenes as participants perceived “similar” phosphenes on 66% of trials (Rizzo et al., 2003). But more recent studies indicated that phosphenes tend to merge in nontrivial ways. For instance, Wilke et al. (2011b) highlighted the importance of electric crosstalk between electrodes in determining the response to simultaneous stimulation of multiple electrodes. Horsager et al. (2011) found that elicited percepts were affected by other stimulating electrodes (even after temporally staggering pulses to remove electric field interactions) and demonstrated a linear combination of threshold currents for simultaneous stimulation. Using a suprachoroidal prosthesis, Sinclair et al. (2016) found that bipolar electrode configurations produced percepts that were similar in appearance to the summation of the phosphenes that were elicited from the two individual electrodes using a monopolar configuration. Most recently, Yücet al. (2022) identified several factors that might limit the spatial resolution of prosthetic vision, which included retinal damage, electrode-retina distance, and the inadvertent stimulation of nerve fiber bundles. To avoid electric crosstalk and aid the perceptual merging of multi-electrode phosphenes, some researchers (Beauchamp et al., 2020; Oswalt et al., 2021; Christie et al., 2022) considered sequential stimulation paradigms. However, sequential stimulation does not always lead to perceptually intelligible forms or objects; often participants are only able to trace an outline of the perceived shape, and their interpretation of the shape relies heavily on this basic outline (Christie et al., 2022). Therefore, understanding how multi-electrode stimulation can be leveraged to produce form vision (that is, a fundamental aspect of visual perception that enables humans to recognize spatial patterns and objects) remains an open challenge for the field of visual prosthetics.

Here we aim to study the consistency and predictability of the (presumably fundamental) building blocks of form vision: the percepts elicited by single- and paired-electrode stimulation. While single-electrode stimulation is relatively well understood (Nanduri et al., 2008; Sinclair et al., 2016; Luo et al., 2016; Beyeler et al., 2019b; Granley and Beyeler, 2021), it remains to be demonstrated whether this knowledge can be extended to predict phosphene appearance elicited by paired-electrode stimulation. Specifically, the axon map model (Beyeler et al., 2019b; Granley and Beyeler, 2021) predicts that the probability of seeing two phosphenes should increase with increasing distance between their axon bundles (as opposed to distance on the retinal surface), but no empirical studies have validated this hypothesis. Moreover, recent computational models of prosthetic vision assume linear summation of phosphenes (Spencer et al., 2019; de Ruyter van Steveninck et al., 2022), but this has yet to be demonstrated empirically. Therefore, to assess whether phosphenes sum linearly and to determine which neuroanatomical and stimulus parameters may be predictive of paired-phosphene appearance, we retrospectively analyzed an extensive psychophysical dataset collected with the help of three Argus II users.

## 2. Methods

### 2.1. Participants

This study involved three blind participants (one female and two male) with severe RP, ranging from 41 to 70 years in age at implantation (Table 1). Participants were chronically implanted with the Argus II Retinal Prosthesis System as part of an interventional feasibility trial (clinicaltrials.gov NCT00407602; completed). All psychophysical experiments were carried out at least six months after device implantation. The study was approved by the Institutional Review Board (IRB) at each participant’s clinical site and was conducted under the tenets of the Declaration of Helsinki. Informed consent was obtained from the participants after explanation of the nature and possible consequences of the study.

**Table 1:** Participant details: sex (M: male, F: female), preoperative visual acuity (VA) categorized as either bare light perception (BLP) or no light perception (NLP), the age range at implantation, and the number of years that participants had been blind prior to implant surgery (self-reported). Years blind for Participant 1 was unknown due to gradual loss of vision.

| Participant ID | Sex | Pre-op VA | Age range at surgery | Years blind |
| --- | --- | --- | --- | --- |
| 1 | M | NLP | 61-70 | ? |
| 2 | F | NLP | 41-50 | 11-20 |
| 3 | M | BLP | 41-50 | 21-30 |

Due to their geographic location, the participants were not directly examined by the authors of this study. Instead, initial experimental procedures were sent to the clinical site, and trained field clinical engineers performed the experiments as specified. Raw collected data was then sent to the authors for subsequent analysis.

### 2.2. Stimuli

Argus II consists of a 6 *×* 10 grid of platinum disc electrodes, each 200 µm in diameter, subtending 0.7^*°*^ of visual angle (Luo and da Cruz, 2016). Electrodes were spaced 575 µm apart. In day-to-day use, an external component is worn by the user, consisting of a small camera and transmitter mounted on a pair of glasses. The camera captures video and sends the information to the visual processing unit (VPU), which converts it into pulse trains using pre-specific image processing techniques (*camera mode*).

All stimuli described in this study were presented in *direct stimulation* mode, where stimuli were sent from the VPU directly to each electrode, without involving the external camera. Stimuli were charge-balanced, cathodic-first, square-wave pulse trains with 0.45 ms phase duration and 250 ms total stimulus duration. Stimulus amplitudes, frequencies, and the number of stimulated electrodes varied based on the design of each experiment. Stimuli were programmed in Matlab 7 (Mathworks, Inc.) using custom software, and pulse train parameters (i.e., the electrode(s) to be stimulated, current amplitude, pulse width, inter-pulse interval, and overall stimulus duration) were sent directly to the VPU, which then sent the stimulus commands to the internal portion of the implant using an inductive coil link. The implanted receiver wirelessly received these data and sent the signals to the electrode array via a small cable.

### 2.3. Perceptual thresholds

Perceptual thresholds for individual electrodes were measured using an adaptive yes/no procedure. Custom software was utilized to measure perceptual thresholds on each electrode through a hybrid method combining an adaptive staircase and constant stimuli approach, using charge-balanced, biphasic 20 Hz pulse trains (de Balthasar et al., 2008). The experiment involved five sessions, where each electrode was tested 12 times, interspersed with 32 catch trials across sessions to assess the false alarm rate, with stimulus amplitudes adjusted based on a Weibull function fit to current data. Data from sessions where the false alarm rate exceeded 20% were deemed unreliable and excluded from the analysis. See Appendix A for a more detailed description of the procedure.

### 2.4. Phosphene drawings

Participants were asked to perform a drawing task upon electrical stimulation of the retina. Participants were comfortably seated in front of a touchscreen monitor whose center was horizontally aligned with the participant’s head. The distance between the participant’s eyes and the monitor was 83.8 cm for Participant 1, 76.2 cm for Participant 2, and 77.5 cm for Participant 3.

Each stimulus was presented in 5–10 trials randomly amongst other stimuli with different frequency and/or amplitude levels. The stimulus frequency ranged from 6 Hz to 120 Hz, and the amplitude was between 1.25 times threshold to 7.5 times threshold. Within each trial, either one or two electrodes were randomly selected and stimulated; if two electrodes were selected, they were stimulated simultaneously. After delivering each stimulus and before moving to the subsequent trial, participants were asked to trace the perceived shape on the touchscreen monitor. The drawing data was recorded and converted into a binary shape data file using Matlab, and stored for future analysis. All psychophysical experiments were carried out by local field clinical engineers at each participating site, and the results were forwarded to the authors.

This yielded 3587 phosphene drawings across three participants. To make the collected phosphene drawings amenable to automated image analysis, we manually inspected all drawings (see Appendix B for details) to make sure that:

- all drawn contour lines were closed (e.g., when drawing a circle, the starting point of the drawing must touch the endpoint);
- small specs (i.e., phosphene with size smaller than 10 pixels) that appeared in less than 50% of trials for a particular electrode were not counted as additional phosphenes.

As part of this procedure, 13 drawings were removed. The remaining 3574 drawings (2717 single-electrode drawings and 857 paired-electrode drawings; see Table 2) were prepared for statistical analysis (explained in Section 2.8).

**Table 2:** The number of drawings for each participant under single and paired-electrode stimulation, categorized by different amplitude levels (upper) or frequency levels (lower).

| Participant | Amp (×Th) | Single-Electrode | | | | | | | |
| --- | --- | --- | --- | --- | --- | --- | --- | --- | --- |
|  |  | Freq (Hz) |  |  |  |  |  |  |  |
|  |  | 6 | 20 | 24 | 30 | 40 | 60 | 120 |  |
| 1 | 1.5 | 40 | 325 | - | - | - | 20 | 20 |  |
| 2 | 1.5 | 40 | 62 | - | - | 20 | 20 | 20 |  |
| 3 | 1.5 | 40 | 101 | - | - | - | - | - |  |
| 3 | 1.25 | 56 | 80 | 17 | 20 | 20 | 74 | 40 |  |

| Participant | Freq (Hz) | Single-Electrode |  |  |  |  |  |  |  | Paired-Electrode |  |  |  |
| --- | --- | --- | --- | --- | --- | --- | --- | --- | --- | --- | --- | --- | --- |
| | | Amp (×Th) | | | | | | | | Amp (×Th) | | | |
|  |  | 1.25 | 1.5 | 2 | 3 | 4 | 5 | 7.5 |  | 1.25 | 1.5 | 2 | 4 |
| 1 | 20 | 131 | 325 | 374 | 20 | - | 60 | 20 |  | 60 | 78 | 110 | - |
| 2 | 20 | - | 62 | 485 | 20 | 109 | 20 | 20 |  | - | - | 304 | 80 |
| 3 | 20 | 80 | 101 | 343 | 20 | - | 40 | 20 |  | - | 98 | 127 | - |

Since the validity and reliability of the experiment relied on the ability of our participants to accurately draw the perceived phosphenes, a control task was conducted where participants were asked to feel six different tactile shapes made of felt with a cardboard background, and then draw them on a touchscreen (Beyeler et al., 2019b). As the shape of these tactile targets was known and we asked participants to repeat each drawing five times, we were able to determine each participant’s drawing error and bias. A detailed description of this task can be found in the Appendix S2 of Beyeler et al. (2019b). In short, this control established baseline drawing variability for each participant, against which we could compare electrically elicited phosphene drawing variability to determine the stability of phosphene appearance.

### 2.5. Phosphene shape descriptors

We used the `measure` module of scikit-image (version 0.18.3, https://scikit-image.org) to automatically extract phosphenes (connected regions) and their corresponding centroids from each drawing. Phosphene shape was quantified using four parameter-free shape descriptors commonly used in image processing: area, perimeter, major axis length, and minor axis length (Nanduri et al., 2008). An example is shown in Fig. 1. These descriptors are based on a set of statistical quantities known as *image moments* (Hu, 1962). *For an M × N* pixel grayscale image, *I*(*x, y*), where *x* ∈ [1, *M*] and *y* ∈ [1, *N*], the raw image moments *M*_*ij*_ were calculated as:

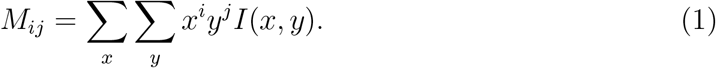

**Figure 1:**
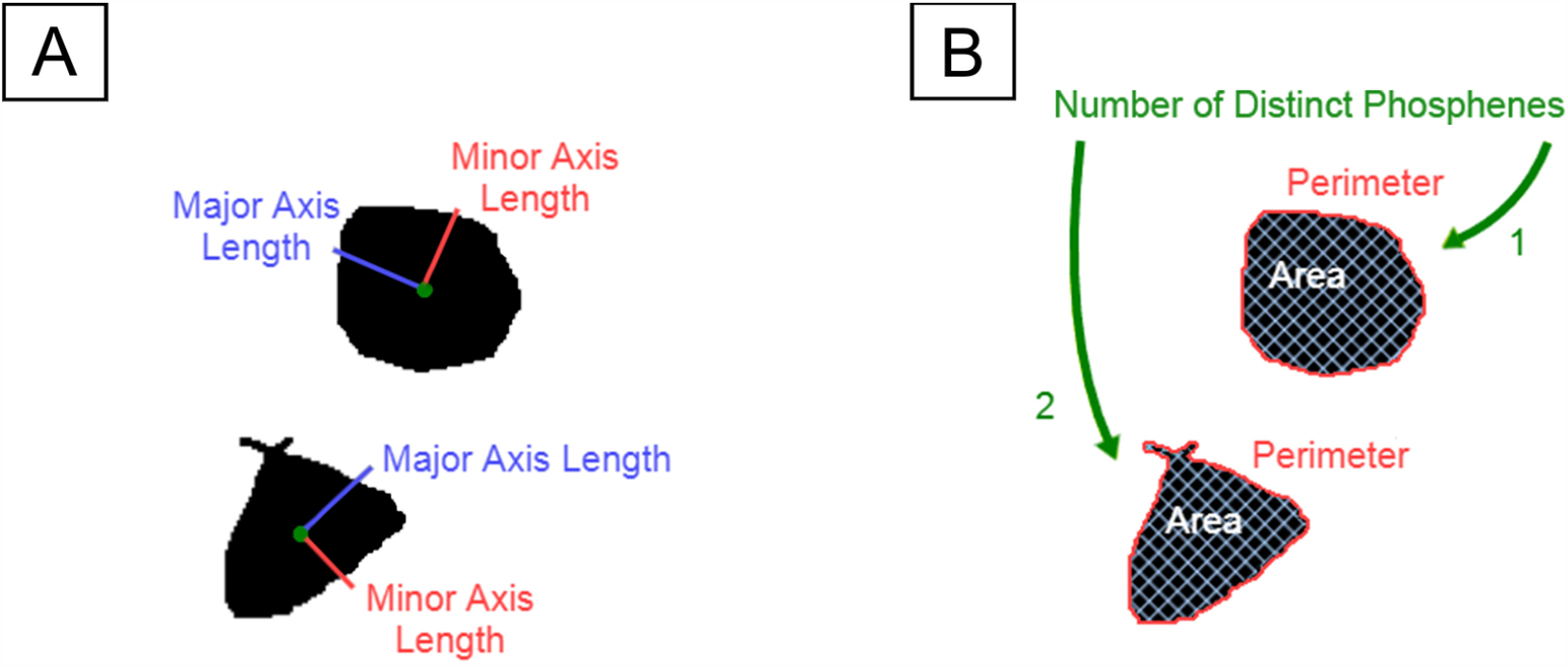
An example of a phosphene drawing and five shape properties of the phosphene. A) Phosphene described by major axis length (red) and minor axis length (blue). B) Phosphene described by area (white), perimeter (red), and the number of distinct regions (green).

Raw image moments were used to compute area (*A* = *M*_00_) and the center of mass 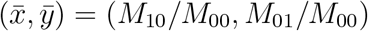 of each phosphene.

Phosphene major/minor axis lengths were calculated from the covariance matrix of the phosphene drawing:

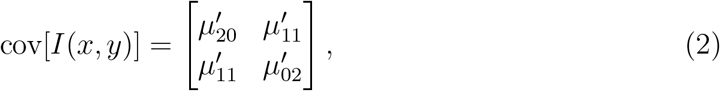

where 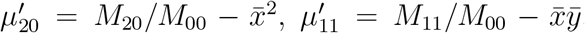, and 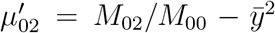. The eigenvectors of this matrix corresponded to the major and minor axes of the image intensity.

Phosphene perimeter was calculated using an algorithm described in Benkrid et al. (2000), which approximates the length of each phosphene’s contour as a line running through the centers of connected border pixels.

The distribution of raw shape descriptors for all participants is given in Appendix C. Phosphene orientation was previously shown to depend mostly on the retinal location of the stimulating electrode (Beyeler et al., 2019b) and was thus excluded from the main analysis. However, the interested reader is referred to Appendix D for the supplemental analysis.

### 2.6. Estimation of electrode-fovea distance and inter-electrode distance

Electrode-fovea distances and inter-electrode distances were estimated using the *pulse2percept* software (Beyeler et al., 2017a). Following Beyeler et al. (2019b), each participant’s implant location was estimated based on the fundus images taken before and after surgery by extracting and analyzing retinal landmarks (e.g., foveal region and optic disc). Image pixels were converted into retinal distances using Argus II inter-electrode spacing information. The implant image was then rotated and transformed such that the raphe fell on the horizontal axis and the fovea was the origin of the new coordination system. The stimulated implant was placed on a simulated map of axonal nerve fiber bundles (Fig. 2), which was modeled based on ophthalmic fundus photographs of 55 sighted participants (Jansonius et al., 2009). Since the fovea is the origin in the stimulated implant’s coordinates, the electrode-fovea distance was measured as the distance between an electrode and the origin.

**Figure 2:**
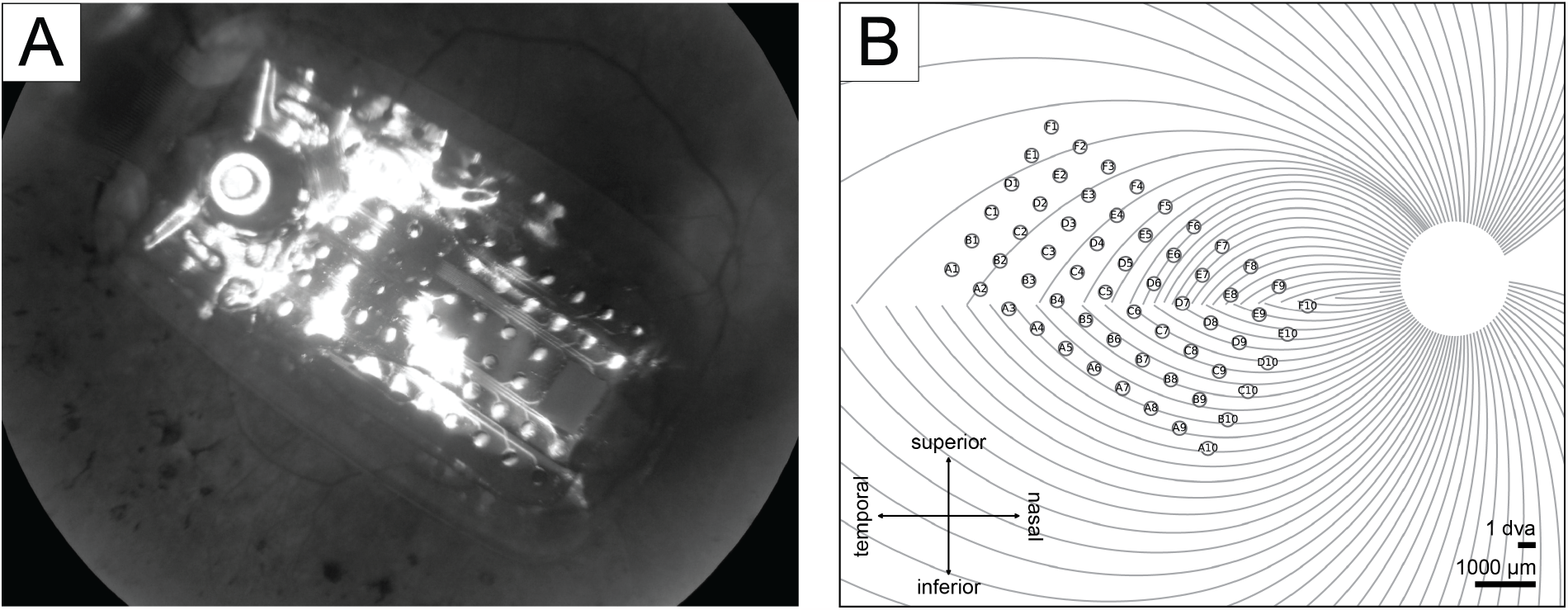
A) Participant 2’s fundus image with Argus II implant placed over the retinal surface. B) Participant 2’s simulated implant placed on the simulated axonal map.

Inter-electrode distance measurements were adapted from Yücel et al. (2022) to investigate the effect of axonal stimulation on perceived phosphene shapes, in which the distance between two electrodes was divided into two, nearly orthogonal components:

- *between-axon* distance (green lines in Fig. 3): the shortest distance between the center of the more nasal electrode to the closest axon of the more temporal electrode;

**Figure 3:**
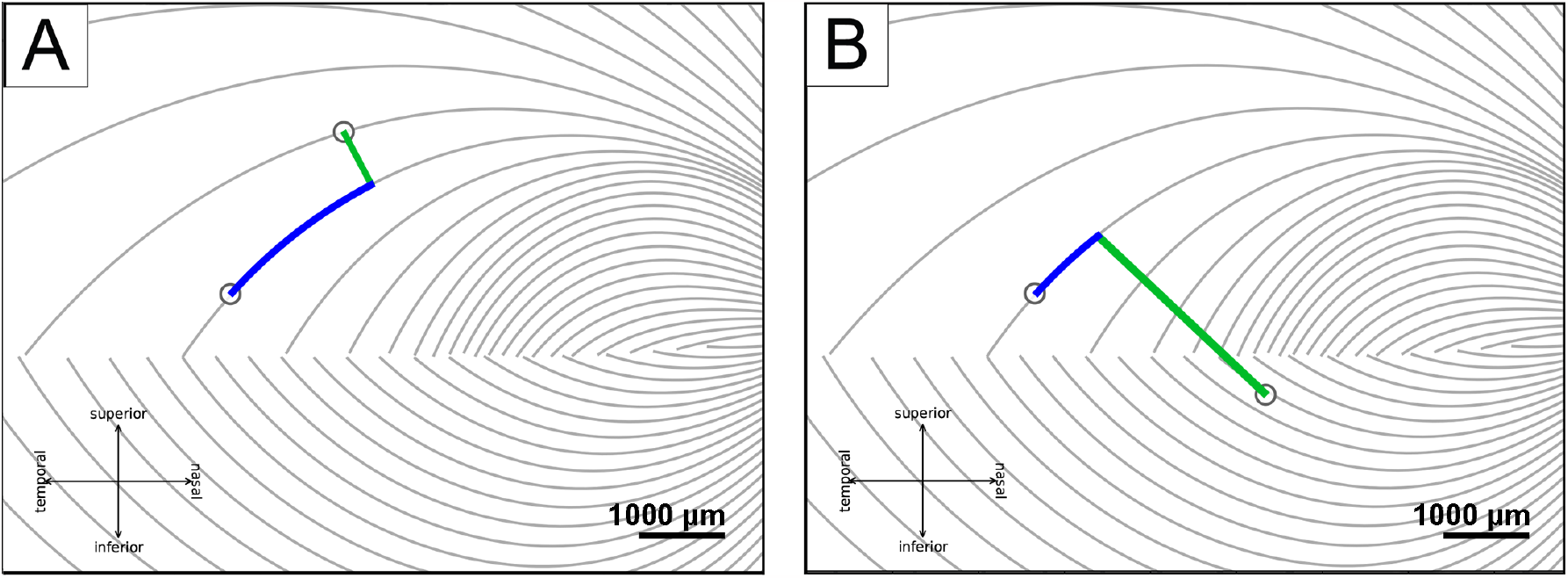
Axonal distances (adapted from Yücel et al., 2022). A) The between-axon distance (green line) and the along-axon distance (blue curve) when two electrodes are on the same side of the raphe, B) and when two electrodes are on different sides.
- *along-axon* distance (blue curves in Fig. 3): the distance from the center of the temporal electrode, along the nasal electrode’s closest axon, up to the point where the nasal electrode’s between-axon line reached the temporal electrode’s axon.

### 2.7. Estimation of electrode-retina distance

Electrode-retina distances were estimated from post-surgical optical coherence tomography (OCT) images collected with either Cirrus HD-OCT (Carl Zeiss Inc) or Topcon 3D-OCT 1000 (Topcon Inc). The SD-OCT scans were obtained 6 months after implantation of Participants 1 and 2, and 13 months after implantation of Participant 3.

When performing OCT scanning, the opaque metal electrodes prevent image acquisition directly underneath the corresponding electrode. However, based on the length of the shadow between the electrode and the retinal surface, it is possible to estimate the electrode-retina distance of that electrode (Ahuja et al., 2013). A single grader manually measured the electrode-retina distance by counting the number of pixels from the center of the shadow on the retinal pigment epithelium to the implant (Fig. 4). These pixel counts were then converted to microns, using the known electrode diameter as a reference to calibrate the pixel-to-micron ratio based on the width of each electrode shadow’s gap in the OCT images. Distances of poorly imaged electrodes were excluded from the dataset.

**Figure 4:**
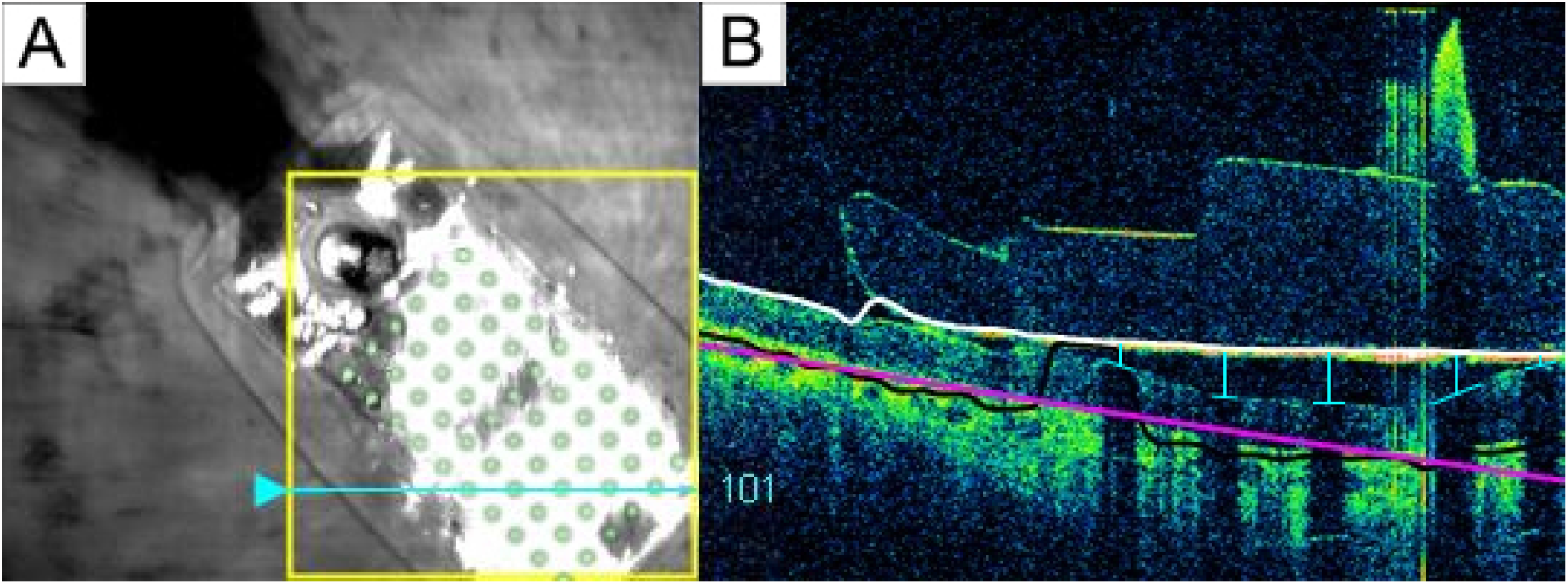
A) Participant 1’s retinal implant fundus image. The cyan arrow marked the current scanning area, and the green electrode array was superimposed onto the original image for better electrode visualization. B) Participant 1’s OCT b-scan. Each electrode-retina distance (vertical blue line) was represented by the length between the center of the shadow on the retinal surface (horizontal blue line) and the implant (white line).

Details about each participant’s estimated electrode-fovea distances and electrode-retina distances are given in Table 3. Welch’s *t*-test was used to compare differences in stimulus and neuroanatomical parameters across participants. There was no statistical difference between the averaged electrode-fovea distance across different participants (for Participants 1 and 2: *t*(29) = 1.529, *p >* .05; for Participants 2 and 3: *t*(29) = 0.114, *p >* .05; for Participants 1 and 3: *t*(29) = *−*1.247, *p >* .05). In terms of electrode-retina distance, Participant 1 had significantly larger values than the other two participants (*t*(29) = 5.776, *p <* .001 and *t*(29) = 5.776, *p <* .001) whose implant was closely attached to the retina.

**Table 3:** Each participant’s number of sampled electrodes, electrode-fovea distance (mean *±* SEM), and electrode-retina distance (mean *±* SEM).

| Participant | Number of included electrodes | Electrode-fovea distance ($\mu\text{m}$) | Electrode-retina distance ($\mu\text{m}$) |
| --- | --- | --- | --- |
| 1 | 30 | $2561.0 \pm 217.5$ | $150.9 \pm 25.5$ |
| 2 | 30 | $2136.2 \pm 173.1$ | 0.0 |
| 3 | 30 | $2168.8 \pm 227.4$ | 0.0 |

### 2.8. Statistical analysis

Data entry and statistical analyses were performed in Python (version 3.8.12, Python Software Foundation). Python package scikit-image (version 0.18.3, https://scikit-image.org) was used for calculating different phosphene shape properties, matplotlib (version 3.5.0, https://matplotlib.org) was used for presenting phosphene drawings and analysis plots, and statsmodels (version 0.13.2, https://statsmodels.org) was used for regression models.

To control for individual drawing bias and variance (Beyeler et al., 2019b) as well as facilitate statistical analysis, we transformed the data as follows:

- All independent variables (i.e., amplitude, frequency, electrode-retina distance, electrode-fovea distance, between-axon distance, and along-axon distance) were standardized across all participants.
- The dependent variables, which describe phosphene shape (i.e., area, perimeter, major/minor axis lengths), were expressed as multiples of the shape descriptors elicited by a “standard” pulse train (amplitude: 2*×* threshold, frequency: 20 Hz). This procedure was performed separately for each participant, but considered drawings from all recorded electrodes of that participant, in order to account for drawing bias and variance. For instance, the area of an individual phosphene was normalized by the phosphene area averaged across all drawings of a particular participant when one of their electrodes was stimulated with the standard pulse.
- Shape descriptors were first extracted from each individual phosphene in each drawing, before they were averaged across trials of the same electrode and stimulus combination, in order to eliminate repeated measures of the same data point. Averaging in this fashion across trials reduced the 3574 drawings to 379 data points (278 single-electrode percepts, 101 paired-electrode percepts).
- Data points that fell more than 2.5 standard deviations away from the mean were considered outliers and were removed from all further analyses. In total, 26 data points were removed from single-electrode analyses, and no data points were removed from paired-electrode analyses. The remaining 353 data points (252 single-electrode percepts, 101 paired-electrode percepts) were included in all analyses.
- Feature descriptors were transformed using a power of 1*/n* to keep the residuals normally distributed. Specifically, we used *n* = 3 for area and *n* = 2 for perimeter, major axis length, and minor axis lengths. All residuals were verified for normality using Quantile-Quantile plots (see Appendix E).

Partial correlation plots for the shape descriptors are given in Appendix E, along with their linear fits.

A series of multiple linear regression and partial correlation analyses were conducted *within* participants (Hou et al., 2023), while linear mixed-effects analyses (with stimulus and neuroanatomical parameters as fixed effects and participants as a random effect) were performed *across* participants.

## 3. Results

### 3.1. Amplitude and frequency modulation affect single-point perception differently

Consistent with the literature on single-electrode phosphene drawings (Nanduri et al., 2008; Luo et al., 2016; Beyeler et al., 2019b), phosphene shape greatly varied across participants and electrodes, but was relatively consistent across trials of a single electrode. Single-electrode stimulation reliably elicited phosphenes in all three participants, who reported seeing a single phosphene on 86.8% of trials, two phosphenes on 13.0% of trials, and three or more phosphenes on the remaining trials.

Fig. 5 shows the mean images for each electrode, obtained by averaging the drawings for each electrode across trials obtained with a particular current amplitude (Fig. 5 *rows*; expressed as a multiple of the threshold current). Mean images were then centered over the corresponding electrode in a schematic of the participant’s implant to reveal the rich repertoire of elicited percepts across electrodes (see Appendix F).

**Figure 5:**
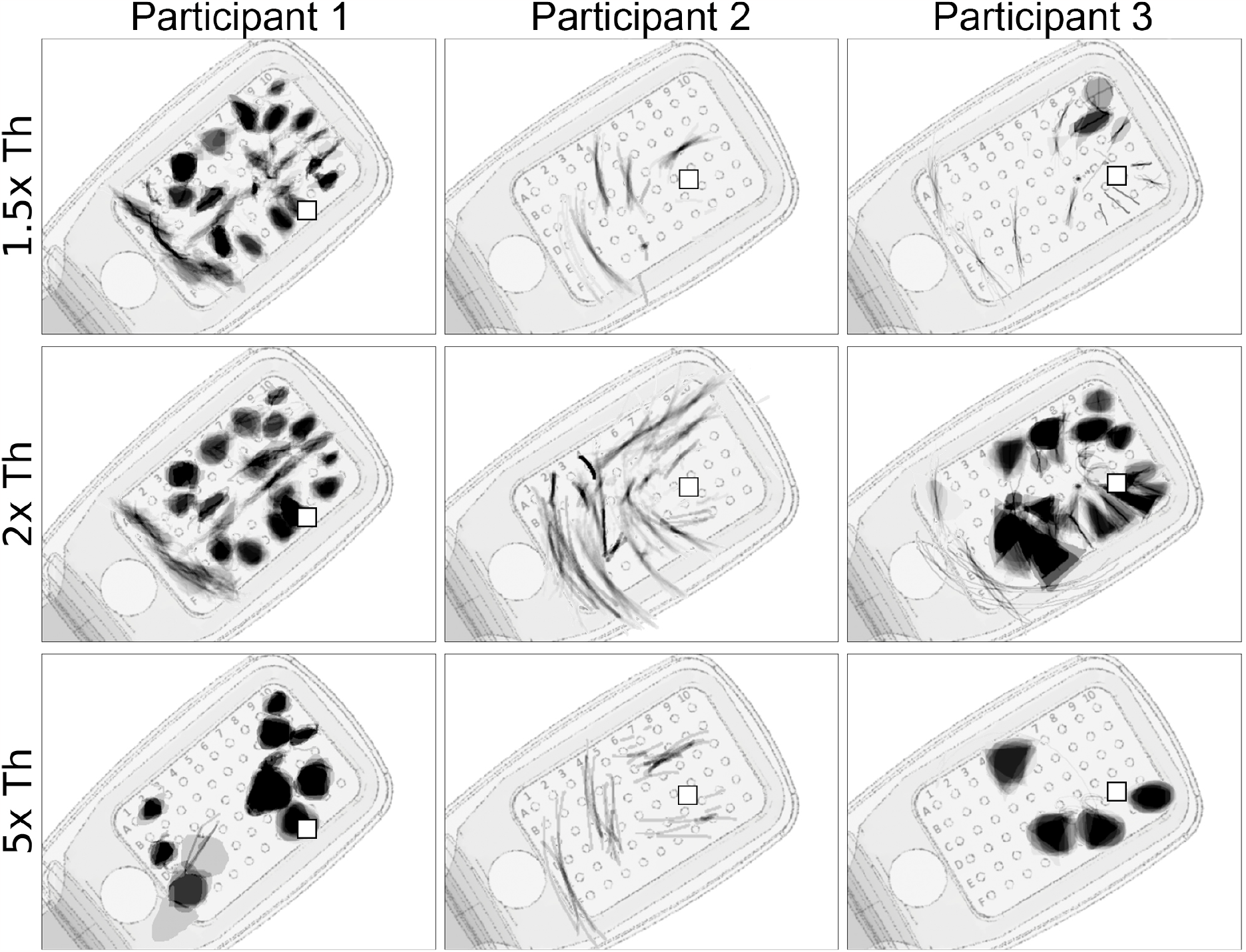
Single-electrode phosphene drawings as a function of stimulus amplitude (*rows*; expressed as multiples of the threshold current). Mean images were obtained by averaging drawings from individual trials aligned at their center of mass (Appendix F). Averaged drawings were then overlaid over the corresponding electrode in a schematic of each participant’s implant (*pulse2percept* 0.8.0.dev0, Beyeler et al., 2017a). Pulse train frequency was 20 Hz for all participants. Squares (□) indicate the estimated location of the fovea.

Whereas Participant 1 mostly drew blobs and wedges, which grew larger as the stimulus amplitude was increased, Participant 2 reported seeing exclusively lines and arcs, which got longer with increasing amplitude. The effect of amplitude on phosphene shape was most apparent for Participant 3, where phosphenes that appeared as lines and arcs near threshold turned into blobs and wedges as amplitude was increased.

First reported by Nanduri et al. (2012), pulse frequency seemed to affect phosphene shape differently than amplitude (Fig. 6). Whereas phosphenes that were located close to the center of vision (denoted by □ in Fig. 6) did not noticeably change in shape, more eccentric phosphenes turned from blobs at 6 Hz to rectangles at 60 Hz (Participant 1), or from short streaks at 6 Hz to orders-of-magnitude longer arcs at 60 Hz (Participant 3).

**Figure 6:**
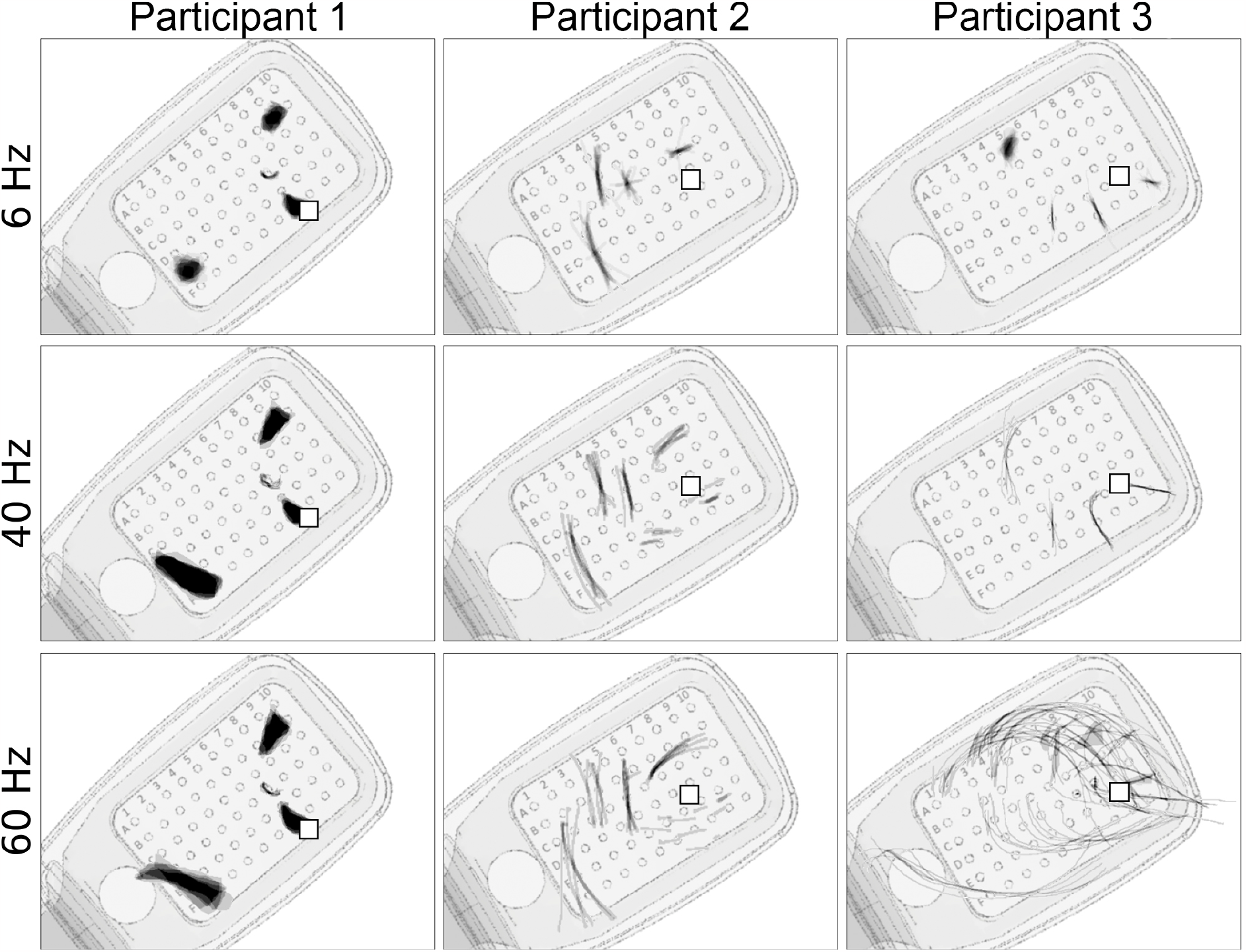
Single-electrode phosphene drawings as a function of pulse train frequency. Mean images were obtained by averaging drawings from individual trials aligned at their center of mass (Appendix F). These averaged drawings were then overlaid over the corresponding electrode in a schematic of each participant’s implant (*pulse2percept* 0.8.0dev; Beyeler et al., 2017a). Shown are only those electrodes for which drawings at all stimulus frequencies were available. Stimulus amplitude was 1.5 times threshold for Subjects 1 and 2, and 1.25 times threshold for Participant 3. Squares (□) indicate the estimated location of the fovea.

### 3.2. Factors affecting phosphene shape during single-electrode stimulation

To more systematically investigate how different stimulus and anatomical parameters affect phosphene shape in single-electrode stimulation, we considered how the four shape descriptors (area, perimeter, major axis length, and minor axis length; see Methods, Section 2.5) could be predicted by different stimulus parameters (i.e., amplitude and frequency) and neuroanatomical parameters (i.e., electrode-retina distance and electrode-fovea distance). To address this, shape descriptor values were first averaged across trials and normalized per participant (see Methods, Section 2.8).

We first performed a multiple linear regression and partial correlation analysis for each participant (top three sections in Table 4), corrected for multiple comparisons with the Bonferroni method. Consistent with Nanduri et al. (2012), we found that stimulus amplitude strongly affected phosphene area in two out of three participants (*p <* .001) and minor axis length (*p <* .001), suggesting that phosphenes tended to get larger with increasing amplitude. However, amplitude did not significantly modulate phosphenes drawn by Participant 2 (also visually evident in Fig. 5). Stimulus frequency had no significant effect on phosphene shape in Participants 1 and 2, but strongly (*β >* .3, *r >* .6) and significantly (*p <* .001) modulated phosphene perimeter, major axis length, and minor axis length in Participant 3.

**Table 4:** Single-electrode phosphene shape predicted by amplitude (Amp), frequency (Freq), electrode-fovea distance (EFD), and electrode-retina distance (ERD; only non-zero for Participant 1). Participant-specific analyses (top three sections) were conducted using a multiple linear regression (*β*: standardized regression coefficient) and partial correlation analysis (*r*: partial correlation coefficient). All-participant analysis (bottom section) was conducted using a linear mixed-effects model fit on all data, with “Participant” as a random factor. Intercepts (not shown) were included in the analysis. The variance inflation factor of all predictors was smaller than 2, suggesting minimal multicollinearity. *N* denotes the number of data points included in each analysis, where each data point represents the mean shape descriptor of the phosphenes elicited with a particular stimulus amplitude and frequency on a particular electrode, averaged across trials. *: *p <* .05, **: *p <* .01, ***: *p <* .001. Significant effects are marked in bold (corrected for multiple comparisons using the Bonferroni method).

|  |  | Area |  | Perimeter |  | Major axis length |  | Minor axis length |  |
| --- | --- | --- | --- | --- | --- | --- | --- | --- | --- |
| | | $\beta$ | $r$ | $\beta$ | $r$ | $\beta$ | $r$ | $\beta$ | $r$ |
| Participant 1<br>($N = 102$) | Amp | **.117\*\*\*** | **.437** | .0333 | .218 | -.00904 | -.0510 | **.132\*\*\*** | **.530** |
|  | Freq | .107 | .257 | .0564 | .221 | .0584 | .194 | .0555 | .156 |
|  | EFD | -.0325 | -.131 | .0443 | .280 | **.0679\*\*\*** | **.353** | -.0425 | -.194 |
|  | ERD | **-.0871\*** | **-.338** | -.0166 | -.110 | -.00497 | -.0279 | -.0650 | -.293 |
| Participant 2<br>($N = 64$) | Amp | .0184 | .125 | .0512 | .242 | .0522 | .242 | .0435 | .241 |
|  | Freq | .0498 | .209 | .104 | .304 | .113 | .323 | .0789 | **.272** |
|  | EFD | **.0874\*\*\*** | **.634** | **.112\*\*\*** | **.601** | **.116\*\*\*** | **.608** | **.0701\*\*\*** | **.482** |
| Participant 3<br>($N = 86$) | Amp | **.255\*\*\*** | **.678** | **.121\*\*** | **.408** | .0290 | .0943 | **.298\*\*\*** | **.575** |
|  | Freq | .0557 | .207 | **.360\*\*\*** | **.813** | **.438\*\*\*** | **.833** | **.304\*\*\*** | **.602** |
|  | EFD | **.0586\*** | **.321** | **.151\*\*\*** | **.665** | **.186\*\*\*** | **.698** | **.104\*\*** | **.367** |
| All Participants<br>($N = 252$) | Amp | **.103\*\*\*** | **.440** | **.0621\*\*\*** | **.297** | .0276 | .118 | **.121\*\*\*** | **.420** |
|  | Freq | **.0400\*** | **.190** | **.156\*\*\*** | **.617** | **.186\*\*\*** | **.624** | **.125\*\*\*** | **.435** |
|  | EFD | **.0426\*** | **.190** | **.107\*\*\*** | **.469** | **.128\*\*\*** | **.479** | **.0581\*\*** | **.209** |

In terms of neuroanatomical parameters, we considered an electrode’s distance to the fovea (i.e., retinal eccentricity) and distance to the retina (i.e., height). Electrode-*retina* distances (labeled “ERD” in Table 4) were non-zero only in Participant 1, where larger ERDs led to smaller phosphenes (*p <* .05). Interestingly, we found that electrode-*fovea* distance (labeled “EFD” in Table 4) significantly modulated shape in all three participants. For Participant 1, more eccentric phosphenes tended to be more elongated (*p <* .001) but not necessarily larger. For Participants 2 and 3, more eccentric phosphenes tended to be larger overall (affecting all shape descriptors with *p <* .05 or smaller).

To determine which of these correlations were general trends that reached significance across all three participants, we also fitted a linear mixed-effects to all data (bottom section in Table 4), corrected for multiple comparisons (Bonferroni), with “Participant” as a random effect. This analysis revealed that larger stimulus amplitudes tended to elicit larger (*p <* .001) and “blobbier” phosphenes (by means of increased minor axis length; *p <* .001). Perhaps driven by Participant 3’s data, increased stimulus frequencies tended to elicit slightly larger and more extended/less compact phosphenes, by means of the overly increased perimeter (*β* = .156, *r* = .617) and major/minor axis lengths (*β >* .12, *r >* .4; *p <* .001). Increasing retinal eccentricity (EFD) had a similar effect, leading to slightly larger and more elongated phosphenes, by means of the overly increased perimeter (*β* = .107, *r* = .469; *p <* .001) and major axis length (*β* = .128, *r* = .479; *p <* .001). Partial correlation plots can be found in Appendix E.2.

### 3.3. Predicting two-point perception from single-point perception

When two electrodes were stimulated simultaneously, participants reported seeing a single phosphene on 53.1% of trials, two phosphenes on 43.4% of trials, and three or more phosphenes on the remaining trials. Three or more phosphenes were generally encountered when single-electrode stimulation itself produced more than one phosphene. Representative examples of phosphene drawings for different electrode pairs are shown in Fig. 7.

**Figure 7:**
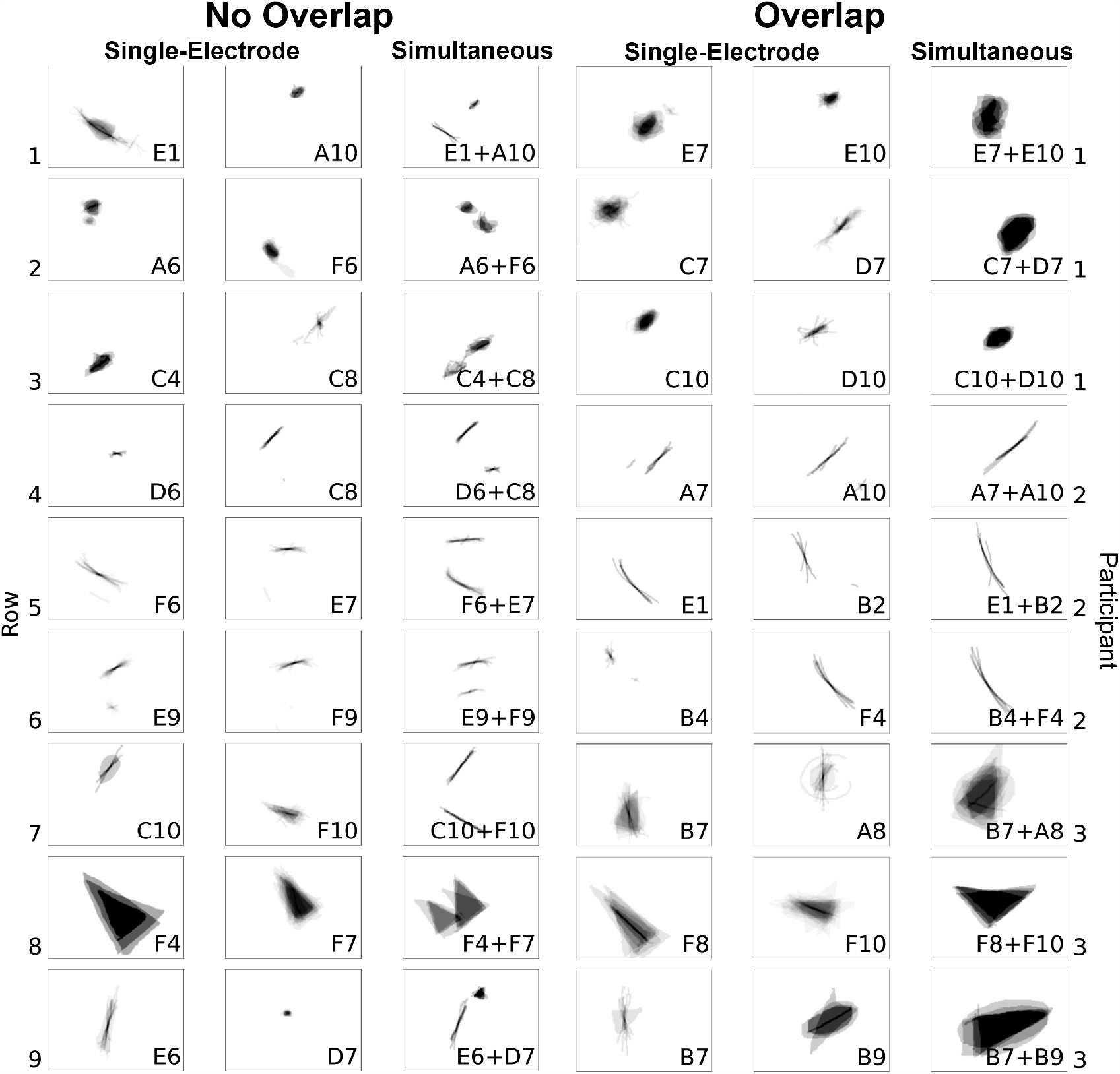
*Left*: Representative examples of single phosphenes combining linearly without overlap during paired-electrode stimulation. *Right*: Representative examples of phosphenes merging and overlapping during paired-electrode stimulation. Mean images were obtained by averaging drawings from individual trials, each phosphene aligned at its trial-averaged center of mass (Appendix F).

When paired-electrode stimulation produced two distinct phosphenes (Fig. 7, *left*), their shape resembled the linear summation of the phosphenes reported during single-electrode stimulation. For instance, as shown in Row 1 of the left panel in Fig. 7, Participant 1 perceived a long arc when electrode E1 was stimulated and an oval when electrode A10 was stimulated. When both E1 and A10 were stimulated concurrently, the resulting phosphene appeared as an arc alongside an oval. Similarly, in Row 9 of the left panel, Participant 3 perceived a tilted line for electrode E6 and a small triangle for electrode D7. Then during the simultaneous stimulation of electrodes E6 and D7, the resulting shape preserved the original form of the individual phosphene shapes. A colored version of this figure that superimposes the outline of the single-electrode phosphenes on the paired-electrode phosphenes is given in Appendix G.

When paired-electrode stimulation produced a single phosphene (Fig. 7, *right*), the phosphenes reported during single-electrode stimulation appeared to merge into a unified shape. For instance, as shown in Row 2 of the right panel in Fig. 7, Participant 1 perceived a blob for electrode C7 and a right-leaning straight line for electrode D7. When both C7 and D7 were stimulated simultaneously, the participant saw a larger blob tilted rightward. Similarly, in Row 6, electrode B4 elicited a small dot, and electrode F4 elicited a long arc; and simultaneous stimulation yielded an arc-shaped phosphene, appearing as a cohesive shape formed by connecting the two individual shapes.

### 3.4. Factors affecting phosphene shape during paired-electrode stimulation

We wondered whether these stimulus and neuroanatomical parameters could also explain the shape of phosphenes elicited by paired-electrode stimulation. As participants would frequently draw multiple phosphenes during paired-electrode stimulation (Fig. 7), we extracted each shape descriptor for each individual phosphene. Then, we summed all phosphenes’ corresponding shape descriptor within each drawing in order to account for the variable number of perceived phosphenes. Finally, we averaged each shape descriptor of each drawing across trials (see Methods, Section 2.8).

The results are shown in Table 5, and partial correlation plots can be found in Appendix E.2. Similar to the single-point results (Table 4), electrode-fovea distance affected phosphene shape in two out of three participants (*p <* .001), generally increasing the perimeter and major axis length of more eccentric phosphenes. However, in contrast to the single-point results, amplitude (i.e., the average amplitude across the two stimulated electrodes) had a less definitive effect on phosphene shape, only increasing the minor axis length (*p <* .05) for Participant 1. Unfortunately, all paired-electrode drawings were collected at 20 Hz, thus stimulus frequency could not be included in the analysis.

**Table 5:** Paired-electrode phosphene shape predicted by amplitude (Amp), electrode-fovea distance (EFD), and electrode-retina distance (ERD; only non-zero for Participant 1). Participant-specific analyses (top three sections) were conducted using a multiple linear regression (*β*: standardized regression coefficient) and partial correlation analysis (*r*: partial correlation coefficient). All-participant analysis (bottom section) was conducted using a linear mixed-effects model fit on all data, with “Participant” as a random factor. Intercepts (not shown) were included in the analysis. The variance inflation factor of all predictors was smaller than 3, suggesting minimal multicollinearity. *N* denotes the number of data points included in each analysis, where each data point represents the mean shape descriptor of the phosphenes elicited with a particular stimulus amplitude on a particular electrode pair, averaged across trials. *: *p <* .05, **: *p <* .01, ***: *p <* .001. Significant effects are marked in bold (corrected for multiple comparisons using the Bonferroni method).

|  |  | Area |  | Perimeter |  | Major axis length |  | Minor axis length |  |
| --- | --- | --- | --- | --- | --- | --- | --- | --- | --- |
| | | $\beta$ | $r$ | $\beta$ | $r$ | $\beta$ | $r$ | $\beta$ | $r$ |
| Participant 1<br>($N = 47$) | Amp | .0841 | .356 | .0332 | .330 | .0104 | .102 | **.0934\*** | **.454** |
|  | EFD | -.0949 | -.248 | .0393 | .240 | .0693 | .378 | -.0895 | -.280 |
|  | ERD | .0446 | .120 | .0232 | .144 | .00886 | .0522 | .0417 | .135 |
| Participant 2<br>($N = 22$) | Amp | .0110 | .0987 | .0112 | .0754 | .0123 | .0827 | -.000242 | -.00188 |
|  | EFD | **.0833\*** | **.599** | **.115\*** | **.613** | **.117\*** | **.619** | .0690 | .472 |
| Participant 3<br>($N = 32$) | Amp | .123 | .375 | .0184 | .156 | -.0271 | -.223 | .166 | .446 |
|  | EFD | .0786 | .250 | .0527 | .412 | **.0652\*** | **.483** | .0171 | .0514 |
| All Participants<br>($N = 101$) | Amp | .0421 | .149 | .00592 | .0643 | -.00954 | -.0314 | .0941 | .169 |
|  | EFD | .0579 | .165 | **.0649\*\*\*** | **.418** | **.0727\*\*\*** | **.459** | .0331 | .0496 |

Naturally, we asked to what extent the phosphene shape elicited by paired-electrode stimulation could be predicted by the phosphene shape elicited during single-electrode stimulation. To answer this question, we conducted a simple linear regression (Table 6) where each shape descriptor from a paired-electrode stimulation trial (e.g., the sum of phosphene areas when Electrodes A and B were simultaneously stimulated) was regressed on the same shape descriptor from a single-electrode stimulation trial (e.g., phosphene area elicited by Electrode A plus phosphene area elicited by Electrode B). In short, we found that each paired-electrode shape descriptor could be predicted by the sum of the two corresponding single-electrode shape descriptors (Table 6; *p <* .001). Across all participants, shape descriptors tended to sum linearly, with the *β* values suggesting that phosphenes elicited by paired-electrode stimulation appeared larger than the average of their single-electrode counterparts, but smaller than their sum.

**Table 6:** Paired-electrode phosphene shape descriptors predicted by the sum of the corresponding single-electrode shape descriptors. Participant-specific analyses (top three rows) were conducted using a simple linear regression (*β*: standardized regression coefficient) and partial correlation analysis (*r*: partial correlation coefficient). All-participant analysis (bottom row) was conducted using a linear mixed-effects model fit on all data, with “Participant” as a random factor. Intercepts were not included in the analysis, because if the value of a predictor (sum of the single-electrode phosphene shapes) was zero, the corresponding value of the dependent variable (the paired-electrode phosphene shape) should also be zero. *N* denotes the number of data points included in each analysis, where each data point represents the mean shape descriptor of the phosphenes elicited on a particular electrode pair, averaged across trials. *: *p <* .05, **: *p <* .01, ***: *p <* .001. Significant effects are marked in bold (Bonferroni-corrected for multiple comparisons).

|  | Area |  | Perimeter |  | Major axis length |  | Minor axis length |  |
| --- | --- | --- | --- | --- | --- | --- | --- | --- |
| | $\beta$ | $r$ | $\beta$ | $r$ | $\beta$ | $r$ | $\beta$ | $r$ |
| Participant 1 ($N = 47$) | **.686\*\*\*** | **.155** | **.593\*\*\*** | **.469** | **.549\*\*\*** | **.670** | **.726\*\*\*** | **.560** |
| Participant 2 ($N = 22$) | **.725\*\*\*** | **.883** | **.726\*\*\*** | **.884** | **.726\*\*\*** | **.883** | **.640\*\*\*** | **.607** |
| Participant 3 ($N = 32$) | **.578\*\*\*** | **.570** | **.558\*\*\*** | **.445** | **.512\*\*\*** | **.396** | **.589\*\*\*** | **.506** |
| All Participants ($N = 101$) | **.653\*\*\*** | **.483** | **.619\*\*\*** | **.649** | **.592\*\*\*** | **.652** | **.692\*\*\*** | **.707** |

### 3.5. Factors affecting the number of perceived phosphenes during paired-electrode stimulation

Yücel et al. (2022) previously demonstrated that the probability of perceiving two distinct phosphenes increases with inter-electrode distance. However, the axon map model (Beyeler et al., 2019b) makes a more nuanced prediction: participants should be more likely to see two distinct phosphenes as the distance between two nerve fiber bundles increases (“between-axon” distance; as opposed to distance on the retinal surface; see Methods). Under this model, paired-electrode stimulation with a short between-axon distance should activate the same nerve fiber bundles and thus lead to a single phosphene, even though the two electrodes may be far apart on the retina.

To test this hypothesis, we split retinal distance into two, almost orthogonal components: “between-axon” distance, which spreads the current radially from the more nasal electrode until it reaches the more temporal electrode’s closest axon, and “along-axon” distance, which walks along the axon from that point until it reaches the more temporal electrode (Fig. 3; this works even for pairs on opposite sides of the raphe). During the preliminary stage of this study, we experimented with a number of similar formulations of splitting these two components, and all of them gave similar results.

Consistent with the axon map model (Beyeler et al., 2019b), we found a significant correlation between the number of perceived phosphenes and the between-axon distance (*p <* .05, Table 7). Along-axon distance, on the other hand, was not significantly correlated with the number of perceived phosphenes (*p >* .05).

**Table 7:** Number of perceived phosphenes predicted by amplitude (Amp), electrode-fovea distance (EFD), electrode-retina distance (ERD), between-axon distance, and along-axon distance. Participant-specific analyses were conducted using multiple linear regression (β: standardized regression coefficient) and partial correlation analysis (*r*: partial correlation coefficient); all-participant analysis was conducted using a linear mixed-effects model fit on all data, with “Participant” as a random factor. Intercepts (not shown) were included in the analysis. The variance inflation factor of all predictors was smaller than 3.1, suggesting minimal multicollinearity. *N* denotes the number of data points included in each analysis, where each data point represents the mean number of perceived phosphenes elicited with a particular stimulus amplitude on a particular electrode pair, averaged across trials. *: *p <* .05, **: *p <* .01, ***: *p <* .001. Significant effects are marked in bold (Bonferroni-corrected for multiple comparisons).

| | Participant 1 ($N = 45$) | | Participant 2 ($N = 9$) | | Participant 3 ($N = 31$) | | All Participants ($N = 85$) | |
| --- | --- | --- | --- | --- | --- | --- | --- | --- |
| | $\beta$ | $r$ | $\beta$ | $r$ | $\beta$ | $r$ | $\beta$ | $r$ |
| Amp | -.0432 | -.125 | — | — | .0606 | .128 | -.00979 | -.0212 |
| EFD | .130 | .220 | **.327\*** | **.875** | -.0947 | -.150 | -.0489 | -.0842 |
| ERD | .156 | .246 | — | — | — | — | — | — |
| Between-Axon Dist. | **.340\*\*** | **.498** | **.374\*** | **.879** | .391 | .399 | **.233\*** | **.327** |
| Along-Axon Dist. | .108 | .207 | .0771 | .366 | -.292 | -.362 | -.0149 | -.0333 |

To further demonstrate the predictive power of the between-axon distance, we constructed two sets of models and compared their Akaike Information Criterion (AIC) and Bayesian Information Criterion (BIC) scores:

- Model A: phosphene number = *f* (along-axon distance, additional factors)
- Model B: phosphene number = *f* (between-axon distance, additional factors)

where “additional factors” consisted of the stimulus parameters (e.g., amplitude) and neuroanatomical parameters (e.g., electrode-fovea distances). As is evident in Table 8, we found strong evidence that models relying on between-axon distance (Model B) significantly outperformed models relying on along-axon distance (Model A) in two out of three participants, as well as in the all-participant analysis.

**Table 8:** Predicting the number of perceived phosphenes during paired-electrode stimulation based on the along-axon distance (Model A) and between-axon distance (Model B). Participant-specific analyses (top three sections) were conducted using a simple linear regression; all-participant analysis (bottom section) was conducted using a linear mixed-effects model fit on all data, with “Participant” as a random factor. Performance is measured using the Akaike Information Criterion (AIC) and Bayesian Information Criterion (BIC) scores, where lower scores indicate better performance. A difference in AIC scores (ΔAIC) or BIC scores (ΔBIC) of less than 2 suggests that there is substantial support for both models (i.e., there is no clear preference for one over the other). 2 *≤* ΔAIC *<* 7 or 2 *≤* ΔBIC *<* 6 indicates some evidence against the model with the higher AIC. ΔAIC *≥* 7 or ΔBIC *≥* 6: strong evidence against the model with the higher AIC/BIC. In practical terms, the model with the lower AIC/BIC is significantly better in terms of the balance between goodness of fit and model simplicity (highlighted in bold). Amp: stimulus amplitude; EDR: electrode-fovea distance; ERD: electrode-retina distance.

|  | Model | Parameters | AIC ↓ | BIC ↓ |
| --- | --- | --- | --- | --- |
| Participant 1 | A | Amp + EFD + ERD + along-axon distance | 53.345 | 62.378 |
|  | B | Amp + EFD + ERD + between-axon distance | **43.954** | **52.988** |
| Participant 2 | A | EFD + along-axon distance | 8.846 | 9.438 |
|  | B | EFD + between-axon distance | **-3.220** | **-2.628** |
| Participant 3 | A | Amp + EFD + along-axon distance | 53.388 | 59.124 |
|  | B | Amp + EFD + between-axon distance | **52.345** | **58.081** |
| All participants | A | Amp + EFD + ERD + along-axon distance | 123.676 | 140.775 |
|  | B | Amp + EFD + ERD + between-axon distance | **114.090** | **131.189** |

## 4. Discussion

In this study, we set out to investigate the relationship between single-point and two-point perception of Argus II users. Our results suggest that two-point perception can be predicted by the linear summation of single-point perception, supporting the notion of independent stimulation channels. We also found that the number of perceived phosphenes increased with the between-axon distance of two stimulating electrodes, but not the along-axon distance, thus providing further evidence in support of the axon map model for epiretinal stimulation (Rizzo et al., 2003; Nanduri, 2011; Beyeler et al., 2019b).

These findings contribute to the growing literature on phosphene perception and have important implications for the design of future retinal prostheses, as they may inform the optimal surgical placement of an epiretinal implant (Beyeler et al., 2019a; Bruce and Beyeler, 2022) and constrain AI-based stimulus optimization algorithms (Granley et al., 2022; de Ruyter van Steveninck et al., 2022; Relic et al., 2022).

### 4.1. Phosphene shape is well predicted by stimulus and neuroanatomical parameters

Although a link between neuroanatomical parameters such as electrode-retina distance and perceptual thresholds has been well established in the literature (Mahadevappa et al., 2005; de Balthasar et al., 2008; Ahuja et al., 2013; Pogoncheff et al., 2023), research examining the effect of these parameters on the *shape* of elicited phosphenes has been limited.

We found that phosphenes tended to appear larger and rounder as stimulus amplitude increased (Table 4), which is consistent with previous considerations about the current spread in the retina (de Balthasar et al., 2008; Granley and Beyeler, 2021; Yücel et al., 2022). However, in contrast to Nanduri et al. (2012), we found that stimulus frequency also affected phosphene size (Table 4 and Fig. 6). Perhaps driven by Participant 3’s data, increased stimulus frequencies tended to elicit slightly larger and more extended/less compact phosphenes, by means of overly increased perimeter and major axis length. This relationship between stimulus frequency and phosphene size partially agrees with data from suprachoroidal prostheses, where phosphenes tend to appear thicker or rounder as the stimulation rate increases (Sinclair et al., 2016).

In addition, we found that increased electrode-fovea distance (i.e., retinal eccentricity) led to slightly larger and more elongated phosphenes (Table 4). While more eccentric phosphenes may be elongated along the trajectory of the underlying nerve fiber bundles (Beyeler et al., 2019b), the increased size may be a consequence of ganglion cell receptive fields increasing with eccentricity (Curcio and Allen, 1990). This would agree with psychophysical (Freeman and Simoncelli, 2011; Stingl et al., 2013) and computational considerations (Song et al., 2022), but is an as-of-yet unpublished finding about the appearance of phosphenes elicited by epiretinal implants. Indeed, most phosphene models assume a constant scaling factor between retinal and visual field coordinates (Horsager et al., 2009; Nanduri, 2011; Beyeler et al., 2019b).

### 4.2. Two-point perception is the linear sum of single-point perception

This study demonstrates that the shape of phosphenes elicited by paired-electrode stimulation is well predicted by the linear summation of the shape of their corresponding single-electrode phosphenes (Table 6), supporting the notion of independent channels for phosphene perception. Specifically, *β* values in Table 6 suggest that phosphenes elicited by paired-electrode stimulation were smaller than the sum of their single-electrode counterparts. This finding is partially consistent with Christie et al. (2022), who showed that the phosphene elicited by electrode “quads” was similar to phosphenes elicited by individual electrodes that belonged to the quad, with Wilke et al. (2011a), who showed that single-electrode phosphenes consisting of round dots and lines added up to more complicated patterns when stimulated simultaneously, and with Barry et al. (2020), who reported that multi-electrode percepts in the Orion cortical implant were perceived to be smaller and simpler than the predicted combination of single-electrode phosphene shapes.

The observed linear summation of single-electrode phosphenes provides valuable empirical evidence for future computational model development. Many computational models of prosthetic vision (Chen et al., 2009; Perez-Yus et al., 2017; Sanchez-Garcia et al., 2019; Granley and Beyeler, 2021) assume a linear relationship between stimulus parameters (e.g., amplitude) and phosphene appearance (e.g., brightness). The same is true for the stimulus generation procedure that underlies “video mode” in Argus II. Here we were able to provide empirical evidence for this assumption and detail the factors that affect phosphene appearance during paired-electrode stimulation. These results may thus inform recent AI-based stimulus optimization algorithms (Spencer et al., 2019; de Ruyter van Steveninck et al., 2022; Relic et al., 2022; Granley et al., 2022), which aim to select the optimal stimulation parameters on each electrode based on their predicted effect on phosphene appearance. These insights may also benefit the prediction of phosphene shape in multi-electrode stimulation scenarios (Zrenner et al., 2010; Shivdasani et al., 2017), which aim to arrange individual phosphenes into more complex patterns, with the ultimate goal of producing form vision to support activities such as reading and recognizing objects.

However, it should be noted that multiple phosphene patterns may not automatically group into perceptually intelligible objects (Stingl et al., 2015; Shivdasani et al., 2017; Christie et al., 2022). This “binding problem” (Roelfsema, 2023) also extends to cortical implants. Although a recent study with intracortical electrodes (Chen et al., 2020) showed that macaques could successfully identify the intended shape of a patterned electrical stimulus, human participants implanted with the same technology could not always do that (Fernández et al., 2021). Indeed, human participants implanted with cortical surface electrodes required a dynamic stimulation strategy to allow for perceptual grouping (Beauchamp et al., 2020).

### 4.3. The number of perceived phosphenes depends on the axonal distance in paired-electrode stimulation

While it is not surprising that two electrodes separated by a large retinal distance might produce two distinct phosphenes (Yücel et al., 2022), here we were able to split retinal distance into two (nearly orthogonal) components: between-axon distance, which measures how far the electric current must spread *away from* an axon bundle until it reaches another electrode, and along-axon distance, which measures how far the electric current must spread *along* an axon bundle until it reaches another electrode. We found that models relying on between-axon distance consistently outperformed models relying on along-axon distance when predicting the number of perceived phosphenes (Table 8). This result provides the first computational evidence that paired-electrode epiretinal stimulation is more likely to elicit two distinct phosphenes as the distance between their underlying axon bundles increases (as opposed to retinal distance alone), and provides further evidence in support of the axon map model for epiretinal stimulation (Rizzo et al., 2003; Nanduri, 2011; Beyeler et al., 2019b).

This result has important clinical implications. First, it suggests that a user’s axon map should be considered when deciding on an intraocular surgical placement of the array (Beyeler et al., 2019a), as phosphenes tend to appear elongated in the direction of the nerve fiber bundle that underlies the stimulating electrode (Beyeler et al., 2019b). As the probability of seeing two phosphenes increases with between-axon distance, the largest number of phosphenes should be produced by an implant whose placement maximizes the sum of between-axon distances between all pairs of electrodes in the array. In other words, electrodes that stimulate the same axon bundle (i.e., with zero between-axon distance) are redundant and should therefore be avoided (Beyeler et al., 2017b). Second, rather than arranging their electrodes on a rectangular grid in an attempt to efficiently tile the retinal surface, future epiretinal implants should strive to place every electrode on a different nerve fiber bundle in an attempt to efficiently tile the axon map, which in turn efficiently tiles the visual field (Bruce and Beyeler, 2022). The same principle may be applied to cortical implants, where future devices could arrange electrodes such that they efficiently tile the visual field rather than the cortical surface.

### 4.4. Limitations and future work

Despite the ability of our work to highlight important factors that guide the appearance of phosphenes elicited by retinal implants, it is important to note that our linear analyses cannot identify nonlinear predictors of phosphene shape. Future studies could thus focus on nonlinear (but still explainable) machine learning models (Pogoncheff et al., 2023). In addition, due to data availability, our analyses are currently limited to single- and paired-electrode stimulation in three participants. However, to achieve form vision, it will be important to stimulate more than two electrodes at a time for each participant. Therefore, future studies should investigate whether this linear summation can be extended to more than two electrodes across the Argus II and the broader retinal implant population.

## Data Availability

All data produced will be available online upon acceptance at

https://github.com/bionicvisionlab/2023-ArgusPairs

## Acknowledgments

This work was supported by the National Eye Institute of the National Institutes of Health (R00-EY029329 to MB). The content is solely the responsibility of the authors and does not necessarily represent the official views of the National Institutes of Health.

## Author contributions

JDW and MB designed the study. DN collected the data. YH and JG processed the data. YH and MB analyzed the data and wrote the manuscript. All authors approved the final version of the manuscript.

## Software and data availability

The code to produce the figures and tables is publicly available at https://github.com/bionicvisionlab/2023-ArgusPairs. Data will be made publicly available via Open Science Framework upon acceptance of this article.

## APPENDIX A. Perceptual threshold measurements

Custom software was used to measure the perceptual thresholds on each electrode using a yes-no procedure that was a hybrid between an adaptive staircase and a method of constant stimuli (de Balthasar et al., 2008). Stimuli were charge-balanced, 0.45 ms per phase, cathodic-first, biphasic 20 Hz pulse trains, 250 ms in duration.

The experiment consisted of five sessions, and each electrode was tested 12 times in each session, in random order. 32 catch trials were also interspersed randomly over five sessions to minimize the false alarm rate. Upon stimulation, participants had to report whether they were able to see any phosphenes (detection task). Stimulus amplitudes (for stimulus present trials) for the first block were predetermined (method of constant stimuli). After the first block, a maximum likelihood algorithm fit of a Weibull function to the current data determined the range of the next block of stimulation amplitude values for each electrode. After each block, a confidence interval was acquired for each electrode using a Monte-Carlo simulation based on responses to the previous trials. If the confidence interval for an electrode fell below a pre-set level, trials for that electrode were no longer presented, but trials on the other electrodes continued through a maximum of five blocks.

Results were deemed unreliable if the false alarm rate, determined by the percentage that the participant saw a stimulus during catch trials, was greater than 20%. Data from runs with higher false alarm rates than 20% were removed from the analysis and the runs were repeated.

## Appendix B. Image processing

### Appendix B.1. Phosphene drawings with open contour lines

It was sometimes challenging for our participants to draw fully closed circles, triangles, or wedges. Although a common strategy is to place one finger at the starting location while the other finger traces out the shape (thus simplifying the process of “returning home” and closing the contour), some drawings ended up with open contour lines (Fig. B1). These drawings were identified as follows:

- The drawing was either a hollow circle or triangle and either had a small gap between two endpoints (Panels A and B) or a line that resembled the shape of a circle or triangle (Panels C and D).
- The majority of drawings from the same electrode showed similarly shaped phosphenes which were all filled.

**Figure B1:**
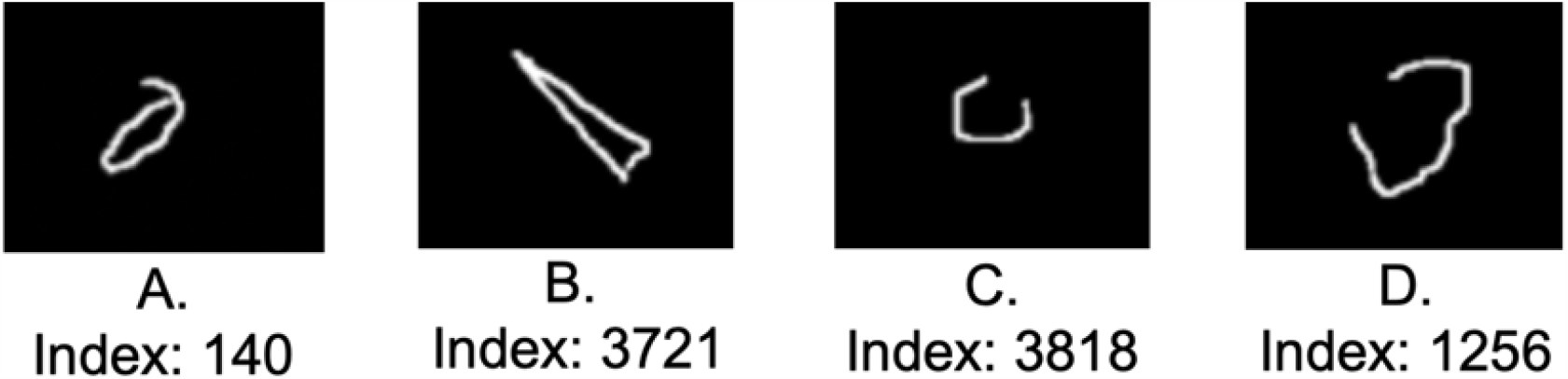
Examples of phosphene drawings with open contour lines.

Based on these criteria, we identified 21 (out of 3587) drawings that needed to be fixed. The data cleaning process involved three steps (Fig. B2):

i. Identify the two endpoints of the drawing (Panel A).
ii. Connect the two endpoints with a 1px-thick line (Panel B).
iii. Fill the area with `scipy.ndimage.binary fill holes()` (Panel C).

Similarly, four phosphenes had small gaps in otherwise smooth contour lines (most likely a tracking issue with the touchscreen). These small artifacts could potentially have grave effects on our phosphene shape analysis, as a gap in the contour line would potentially be judged as two independent, connected regions by the image processing software, thereby accidentally doubling the number of reported phosphenes and halving their reported size.

**Figure B2:**
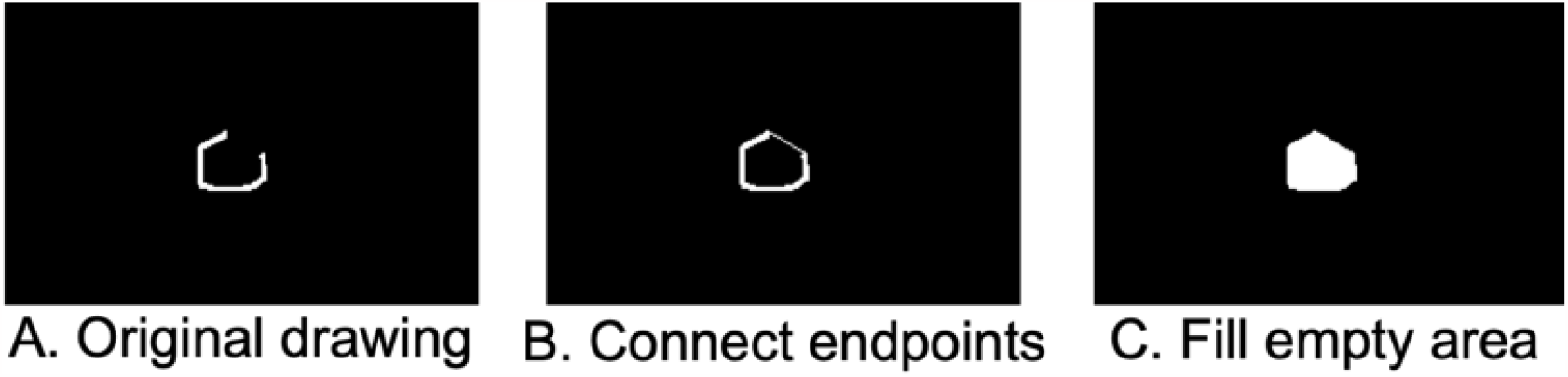
Procedure for fixing phosphene drawings with open contour lines.

Fortunately, we identified only twelve drawings with this issue. To fix them, we manually identified four endpoints of the broken contour line (Fig. B3, Panel A) and connected them (Panel B), then used scipy.ndimage.binary fill holes() to fill the area (Panel C).

**Figure B3:**
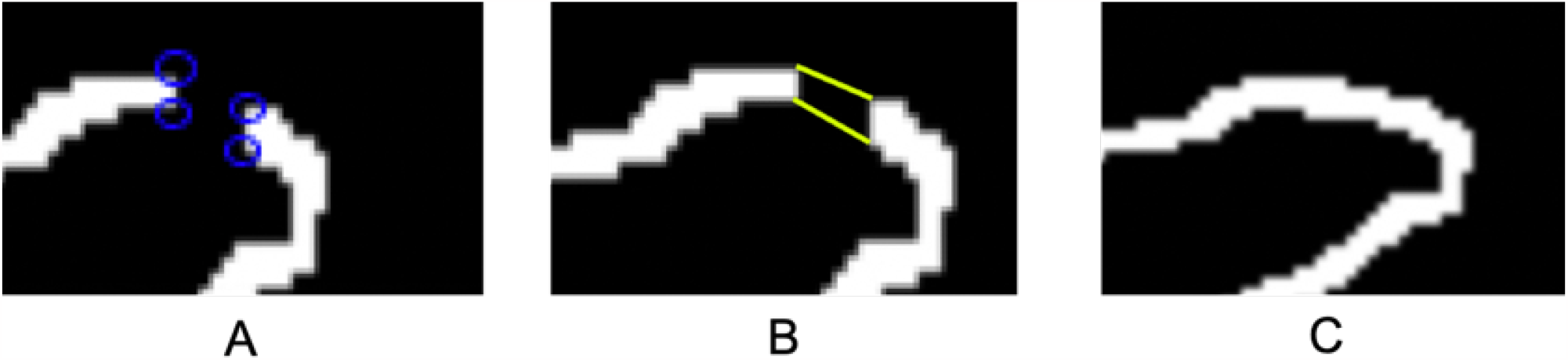
Procedure for fixing phosphene drawings with broken contour lines.

### Appendix B.2. Phosphene drawings with other artifacts

Fourteen phosphene drawings had other artifacts, such as tiny specs (less than 10 pixels in size) that were not part of any other discernible shape, and were subsequently removed (Fig. B4).

**Figure B4:**
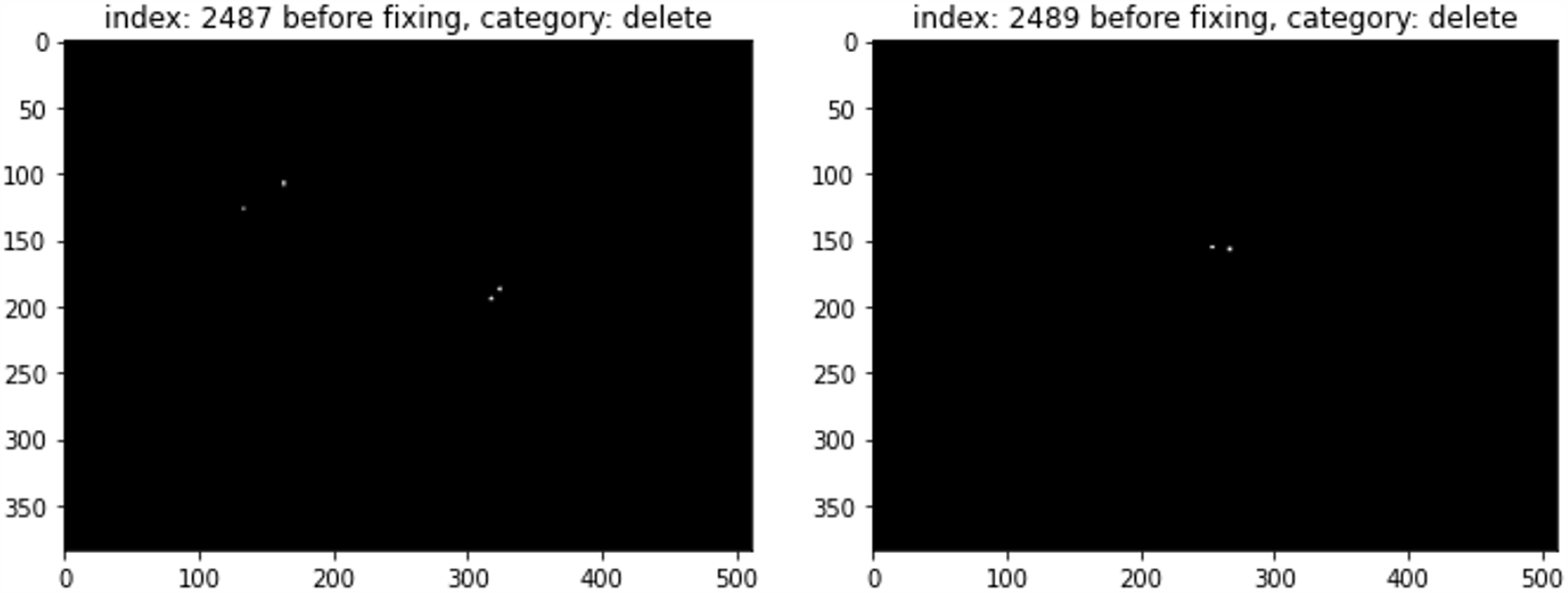
Example phosphene drawings with small specs (artifacts) that were removed from the dataset.

## Appendix C. Distribution of phosphene shape descriptors

Fig. C1 shows the distribution of shape descriptors. Phosphene drawings were more consistent within than across participants (for details, see Nanduri (2011); Beyeler et al. (2019b)). In single-electrode stimulation, Participant 1 tended to draw simple dots, oval and elongated lines varying in length and thickness (Fig. 5; *left column*). However, round or oval shapes never appeared in Participant 2’s drawings, as all phosphenes were curved or straight lines (Fig. 5; *center column*), leading the average phosphene areas, minor axis lengths, and perimeters to be much smaller than those of the other two participants. This was also evident in the boxplots of each participant’s phosphene shapes (Fig. C1; *top row*), because Participant 2’s median area, minor axis length, and perimeter were even smaller than those of other participants’ 25th quantile. The drawings of Participant 3 (which included curved lines, ovals, and triangles) varied dramatically in shape across different electrodes.

Similar tendencies were observed in paired-electrode stimulation (Fig. C1, *bottom row*).

**Figure C1:**
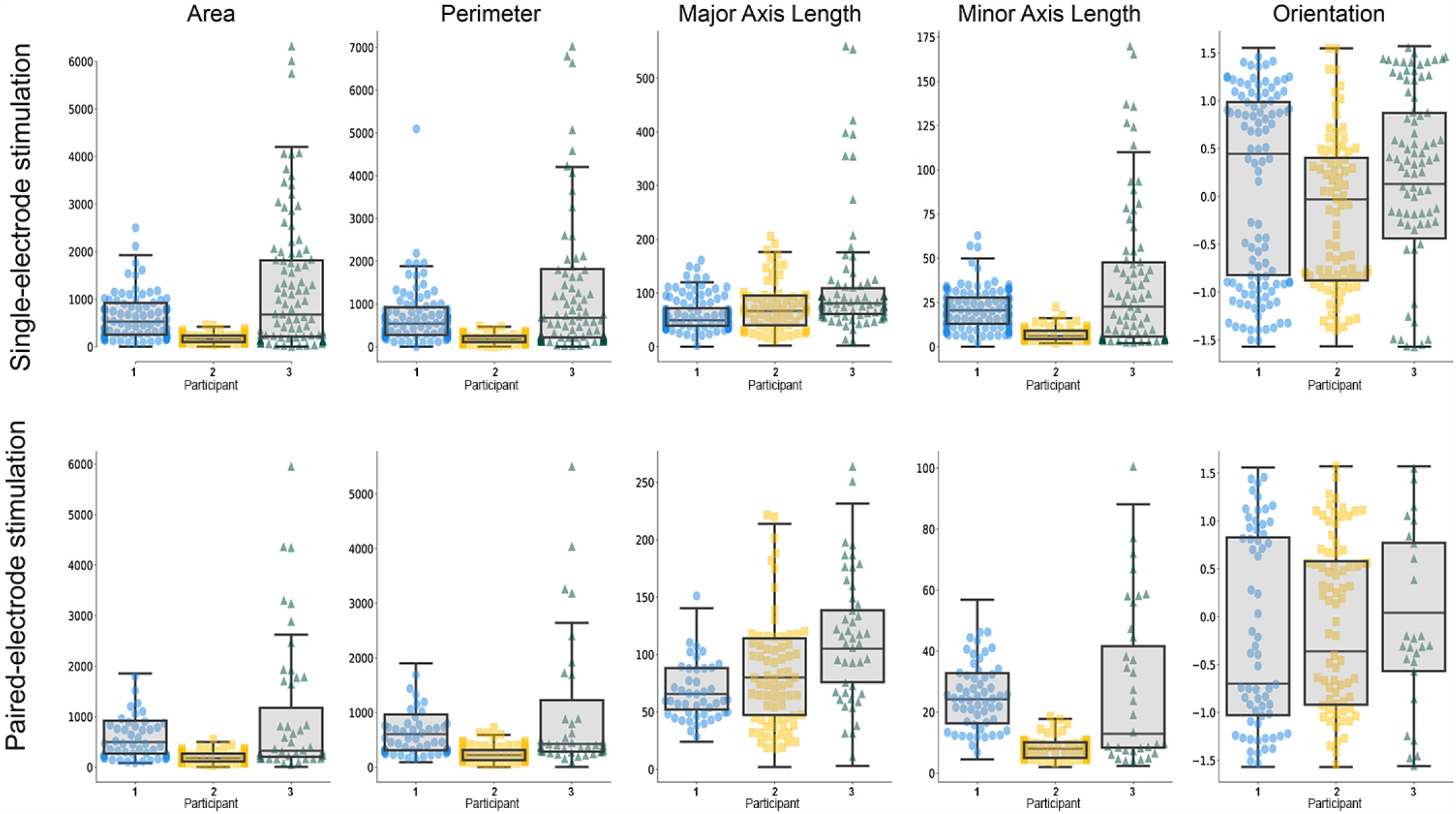
Boxplot of different phosphene shape properties for single-electrode stimulation (top row) and paired-electrode stimulation (bottom row).

## Appendix D. Orientation analysis

Phosphene orientation was also calculated from the covariance matrix of the phosphene drawing:

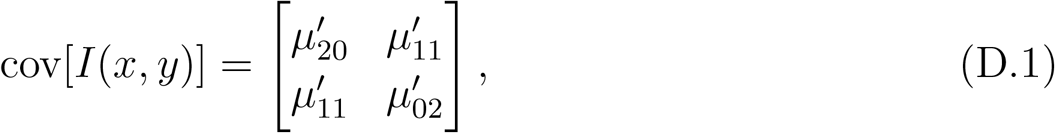

Where 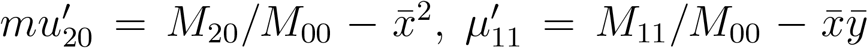, and 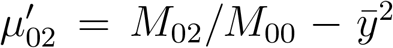. The eigenvectors of this matrix corresponded to the major and minor axes of the image intensity. Phosphene orientation could be extracted from the angle of the eigenvector associated with the largest eigenvalue towards the axis closest to this eigenvector:

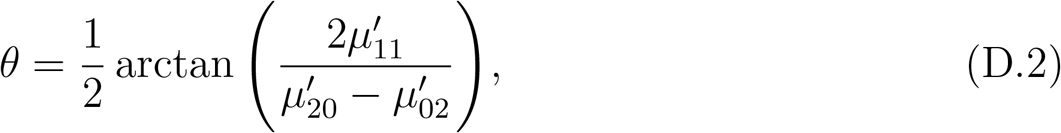

which was valid as long as 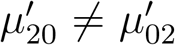, with *θ* ∈ [*−π/*2, *π/*2]. To avoid division by zero, we manually assigned an angle of *θ* = 0 whenever 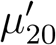 was equal to 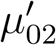.

Consistent with Beyeler et al. (2019b), we found a significant correlation between the orientation of the nerve fiber bundle closest to the stimulating electrode and the orientation of the perceived phosphene (Table D1). This was true for both single-electrode and paired-electrode stimulation experiments except for one of the subjects (*p <* .05; first two modules of Table D1). Moreover, the average of orientations in a paired-electrode stimulus could be predicted by the average orientations of the individual phosphenes measured during single-electrode stimulation (*p <* .001; last module of Table D1), suggesting that the orientation of individual phosphenes did not change much during simultaneous stimulation.

**Table D1:**
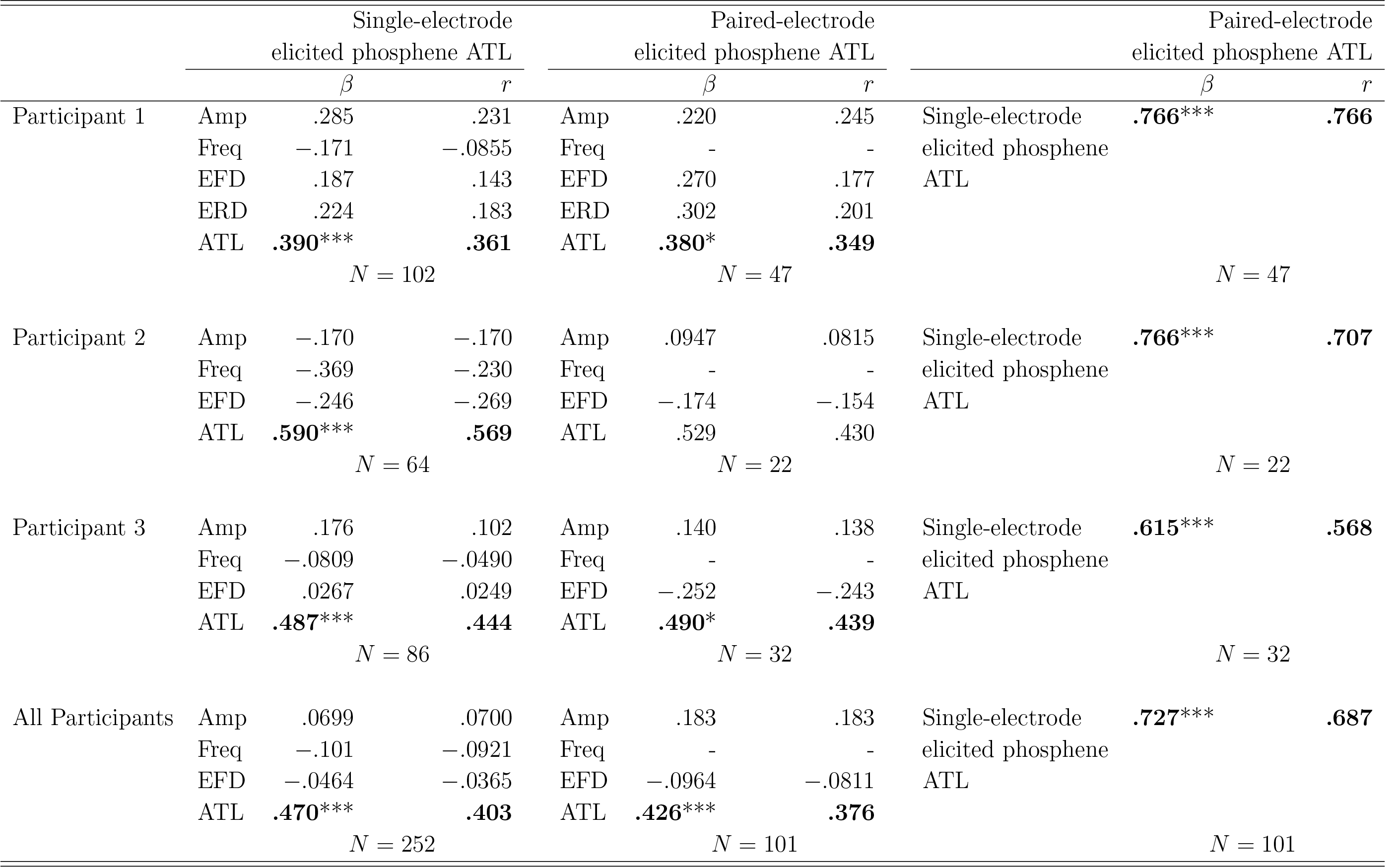
Phosphene numbers predicted by different stimuli and electrode-retina interface properties in single-electrode drawings and paired-de drawings, and phosphene numbers in paired-electrode drawings predicted by phosphene numbers in single-electrode drawings. The ce inflation factor of all predictors was smaller than 3. *Amp*: Amplitude. *Freq*: Frequency. *EFD*: Electrode-Fovea Distance. *ERD*: de-Retina Distance. *ATL*: Axonal Tangential Line. *: *p <* .05, **: *p <* .01, ***: *p <* .001. Significant effects are marked in bold cted for multiple comparisons using the Bonferroni method).

## Appendix E. Statistical analysis

### Appendix E.1. Q-Q plots

We used Q-Q plots with residuals of the multiple linear regression models (per-participant) and linear mixed-effects models (all-participants) to assess the normality of the residuals.

**Figure E1:**
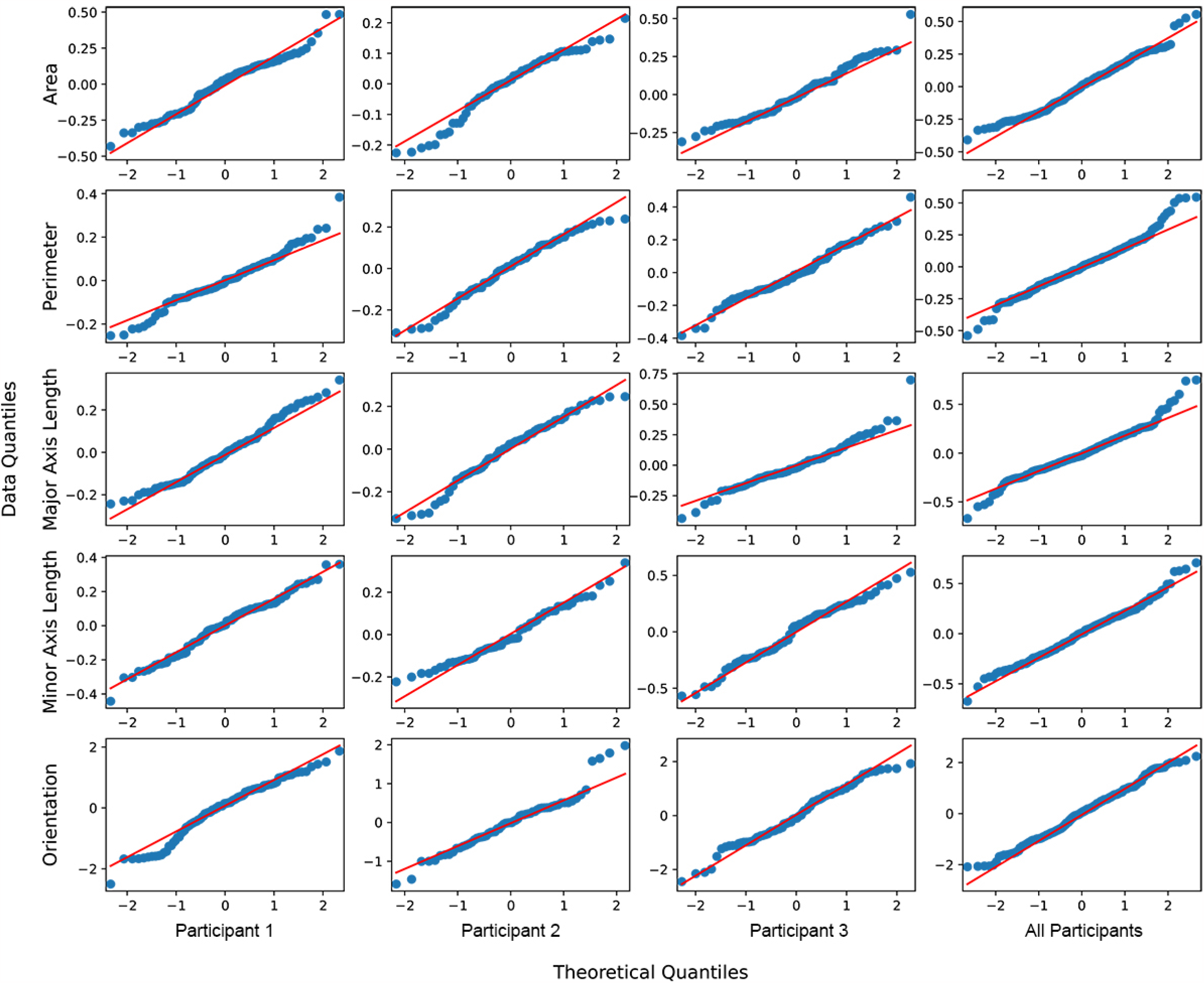
The Q-Q plots of residuals for single-electrode stimulation.

**Figure E2:**
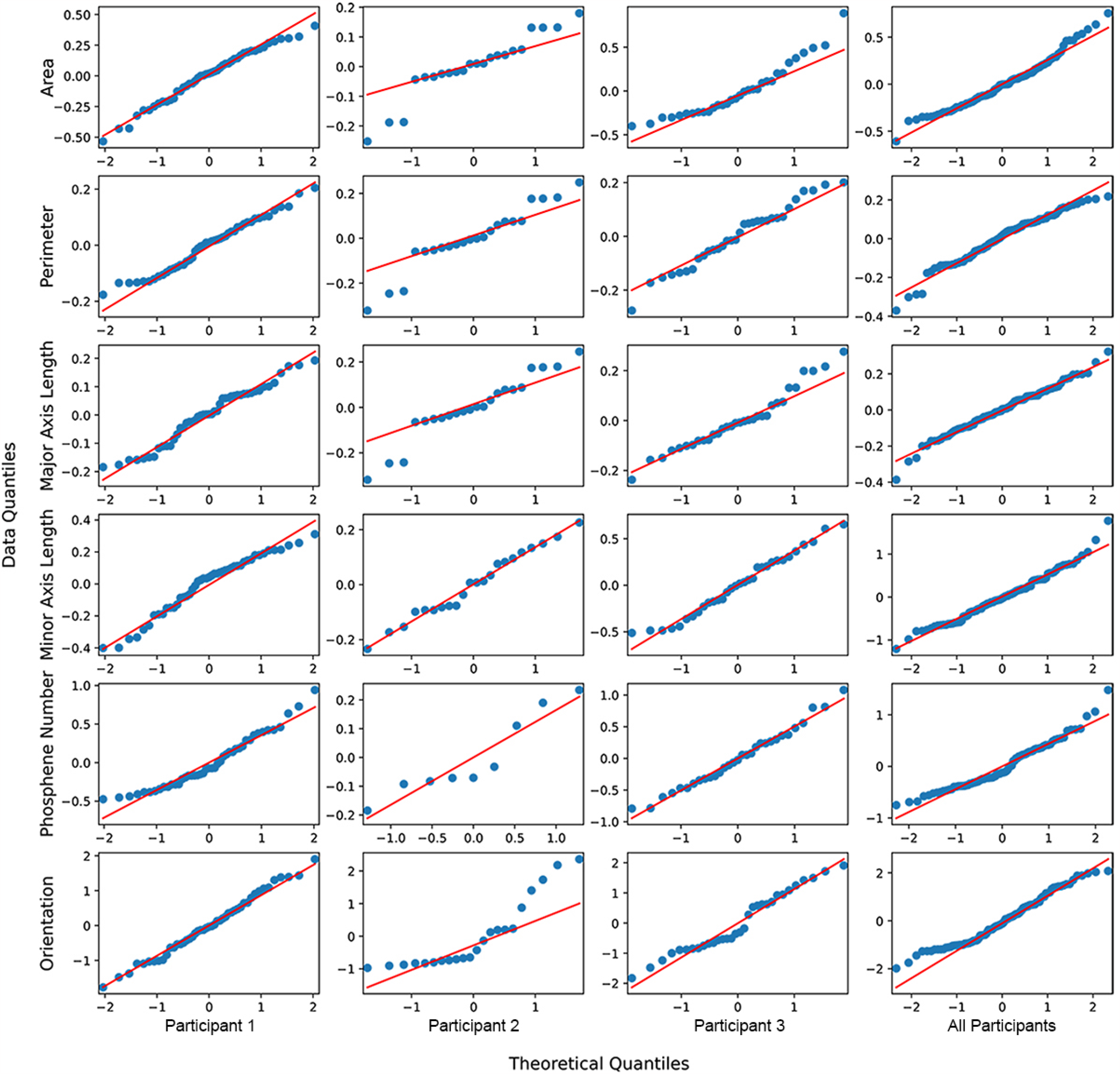
The Q-Q plots of residuals for paired-electrode stimulation.

### Appendix E.2. Partial correlation plots

**Figure E3:**
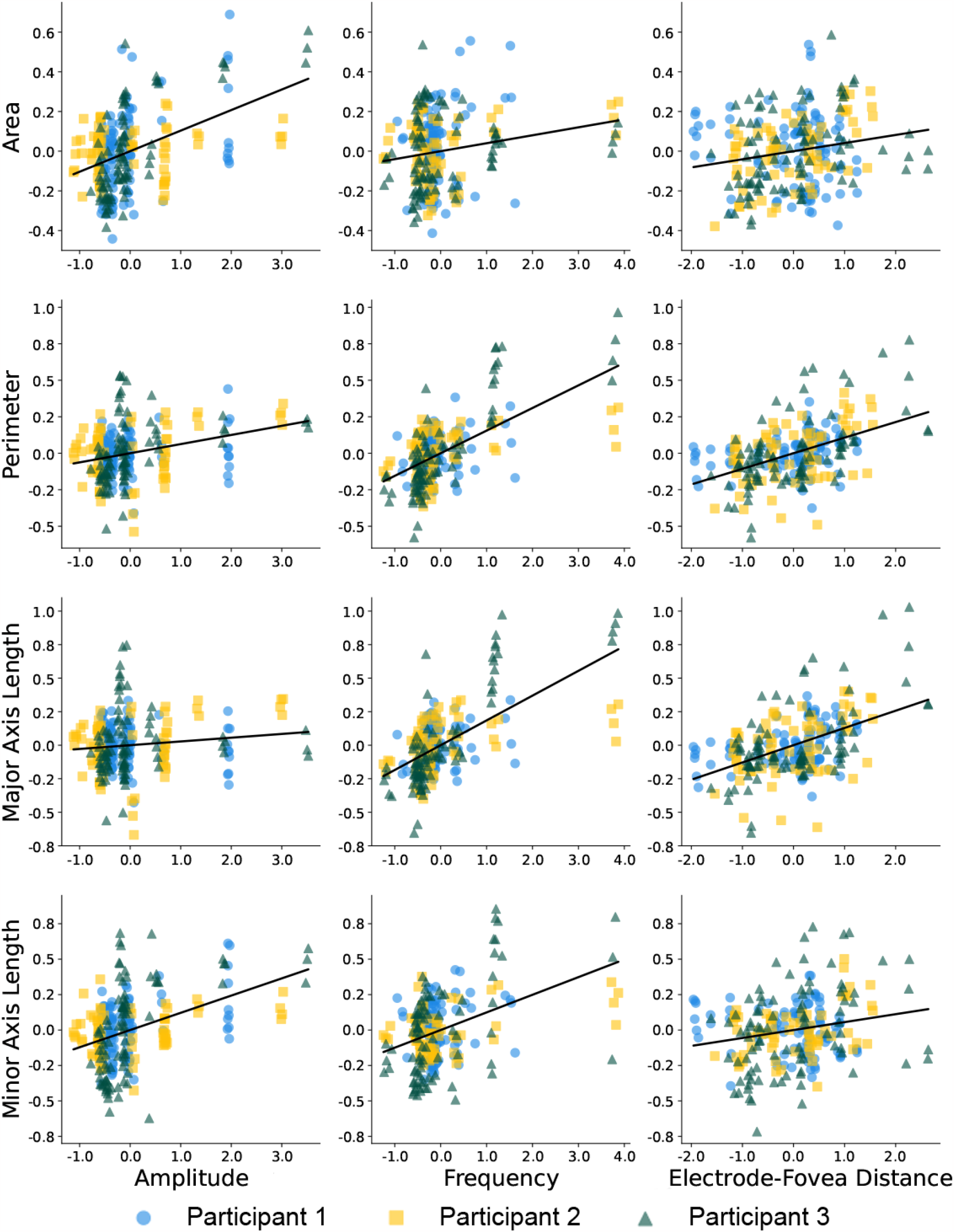
Partial correlation plots of area, perimeter, major axis length, or minor axis length correlated with amplitude, frequency, electrode-fovea distance, and electrode-retina distance across all participants in single-electrode stimulation.

**Figure E4:**
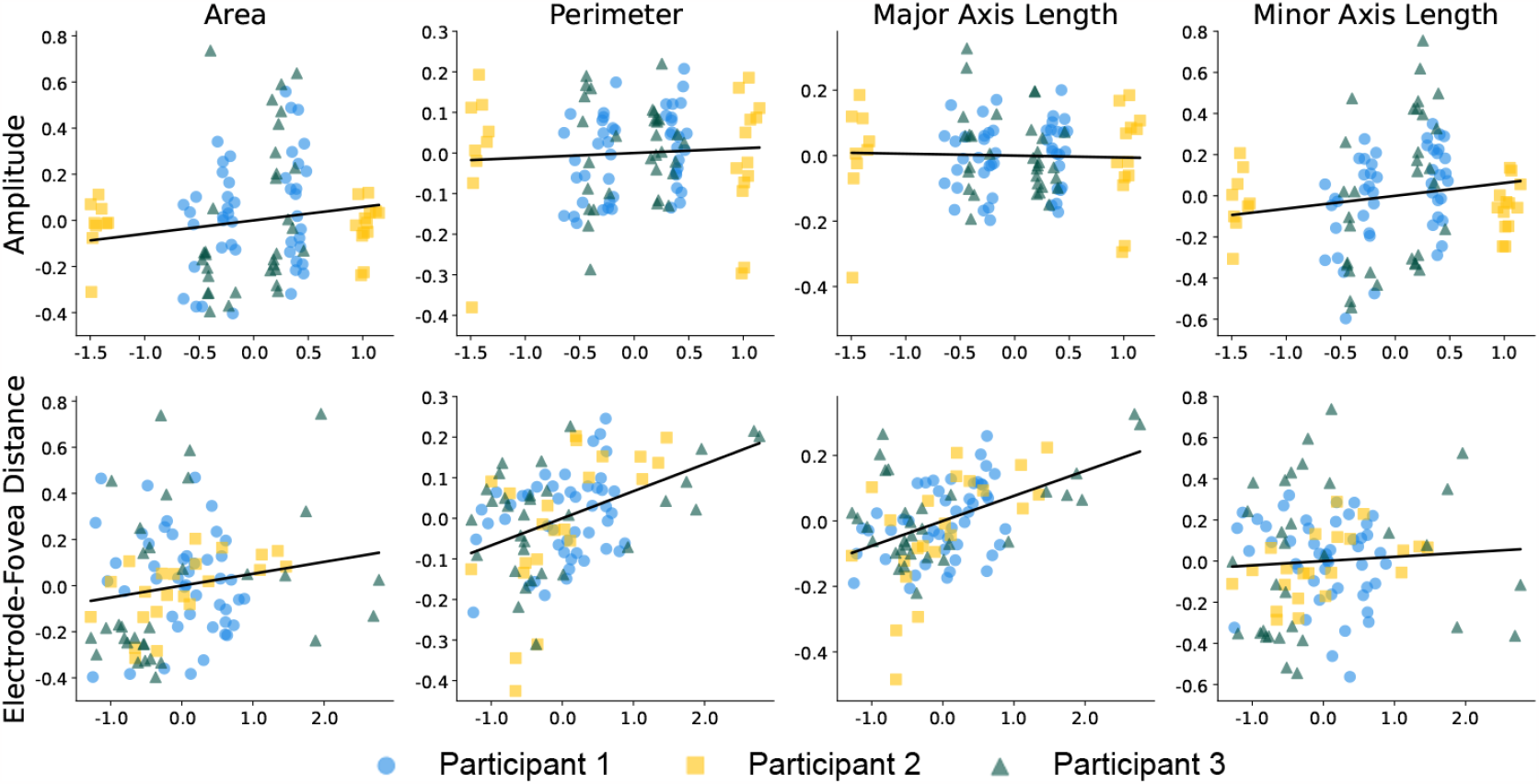
Partial correlation plots of normalized phosphene shape elicited by paired-electrode stimulation, correlated with standardized stimulus amplitude and neuroanatomical parameters (arbitrary units).

**Figure E5:**
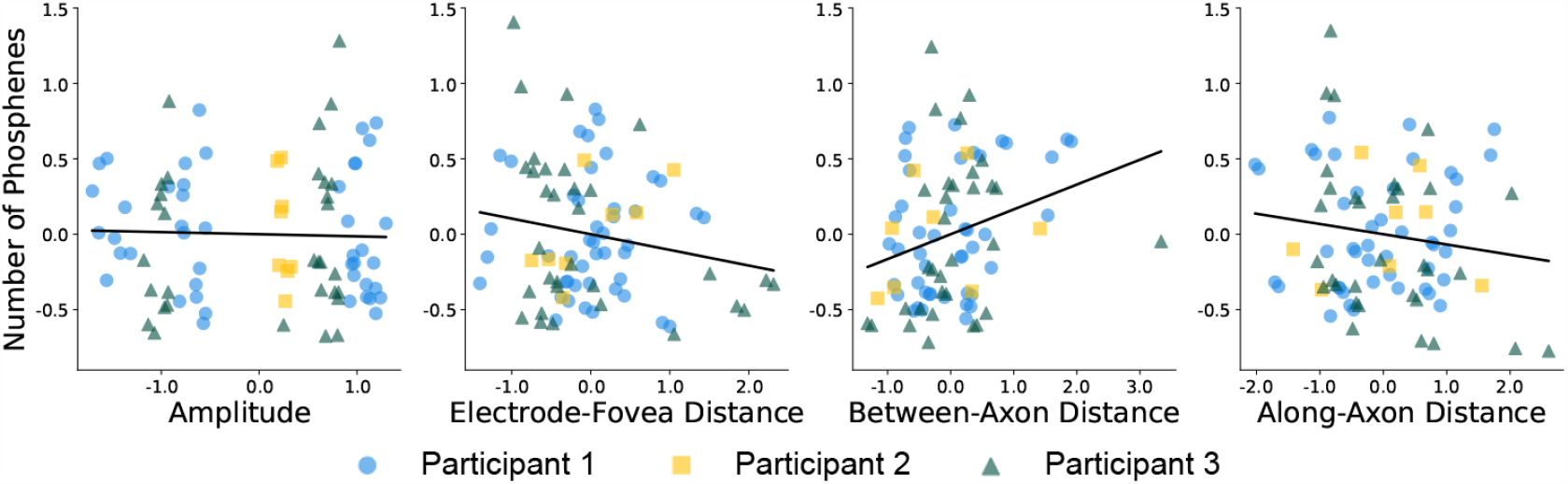
Partial correlation plots of the number of distinct phosphene regions correlated with stimulus parameters and electrode-retina properties across all participants in paired-electrode stimulation.

## Appendix F. Visualizing average phosphenes

To draw mean phosphene shapes for a particular electrode and stimulus (as shown in, for instance, Fig. 5), phosphenes were first aligned by their centroid and averaged, then aligned with the electrode location of the implant schematic (using pulse2percept 0.8.0.dev; Beyeler et al., 2017a). Note that our statistical analyses did not depend on a phosphene’s centroid location, so the following description only serves to produce a meaningful visualization of mean phosphenes.

If all five trial drawings showed exactly one phosphene (86.8% of trials during single-electrode stimulation), alignment and averaging were straightforward. If all five trial drawings showed exactly two phosphenes (43.4% of trials during paired-electrode stimulation), phosphenes were assigned to electrodes by clustering their centroid locations:

- In the drawing of Trial 1, the phosphene whose centroid had the smaller *x* coordinate (if same *x* coordinate: smaller *y*) was labeled as Phosphene A, and the other as Phosphene B.
- In the drawing of Trial 2, the centroid location for each phosphene was compared to the centroids of Trial 1 and assigned to whichever centroid was closest.
- This process was repeated for every phosphene in all subsequent trials until every phosphene was either grouped with Phosphene A or Phosphene B.

If the number of phosphenes varied across trials, a more sophisticated procedure was necessary:

- Find the average centroid location of all single-phosphene drawings (Fig. F1, Panel A). In the paired-phosphene drawings, identify the phosphene that is closest to the average centroid location (“first phosphene centroid”).
- Find all phosphenes belonging to that centroid and average them (Fig. F1, Panel B, “first averaged drawing”).
- Find all other phosphenes that have not been processed yet and average those (Fig. F1, Panel C, “second averaged drawing”).

**Figure F1:**
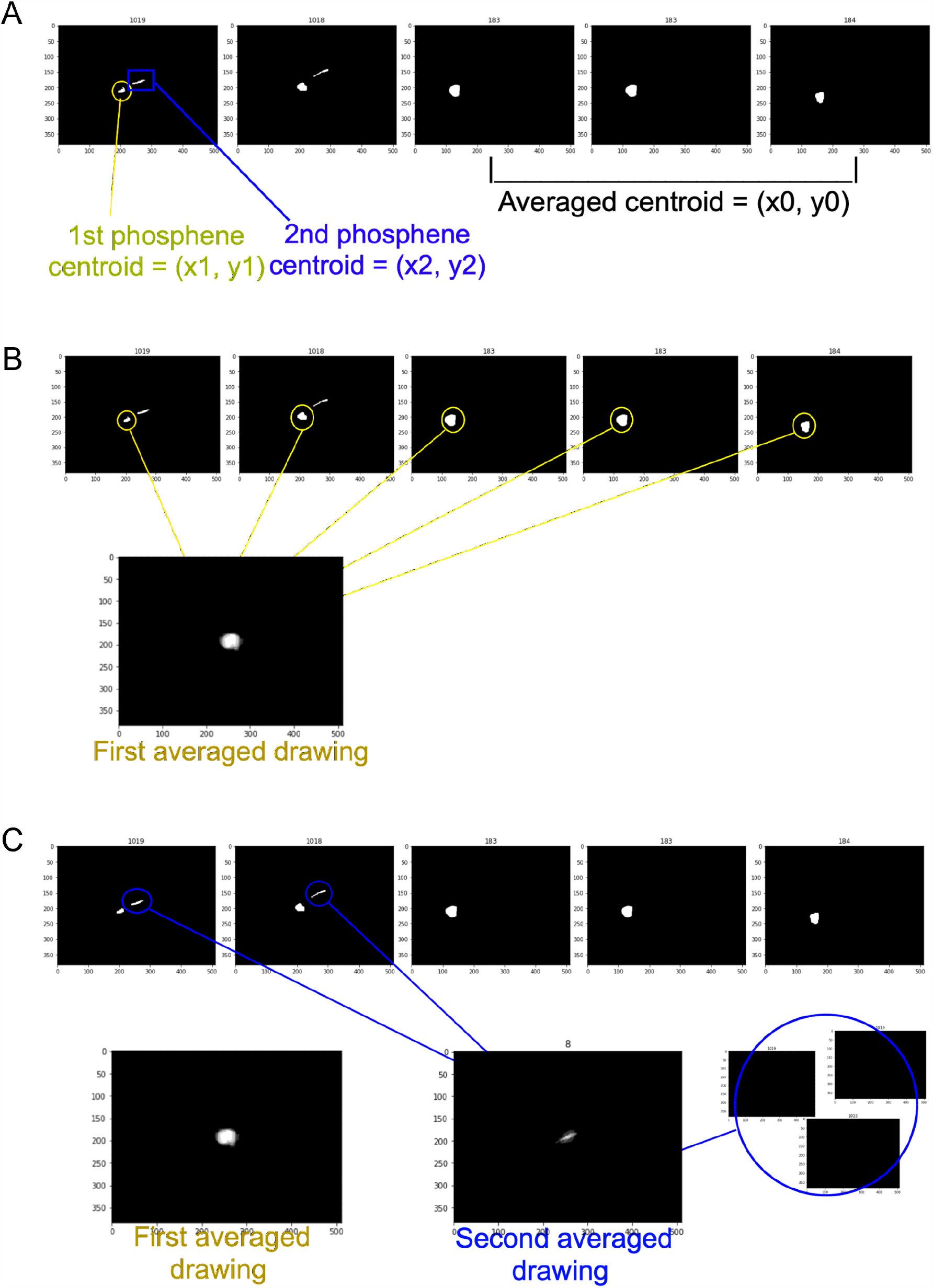
Procedure for stacking drawings when the number of phosphenes differed across trials. Note that this procedure only applies to the visualizations in Figures 5–7, as our statistical analysis did not require phosphene stacking.

## Appendix G. Phosphene superimposition

To better visualize to which extent the phosphenes generated by paired-electrode simultaneous stimulation (Fig. 7) align with the linear combination of phosphenes produced by individual electrode stimulations, we overlaid the outlines of single-electrode phosphenes (first column in Fig. G1) onto the drawings of the phosphenes elicited by paired-electrode stimulation (second column in Fig. G1) for both “No Overlap” and “Overlap” panels.

**Figure G1:**
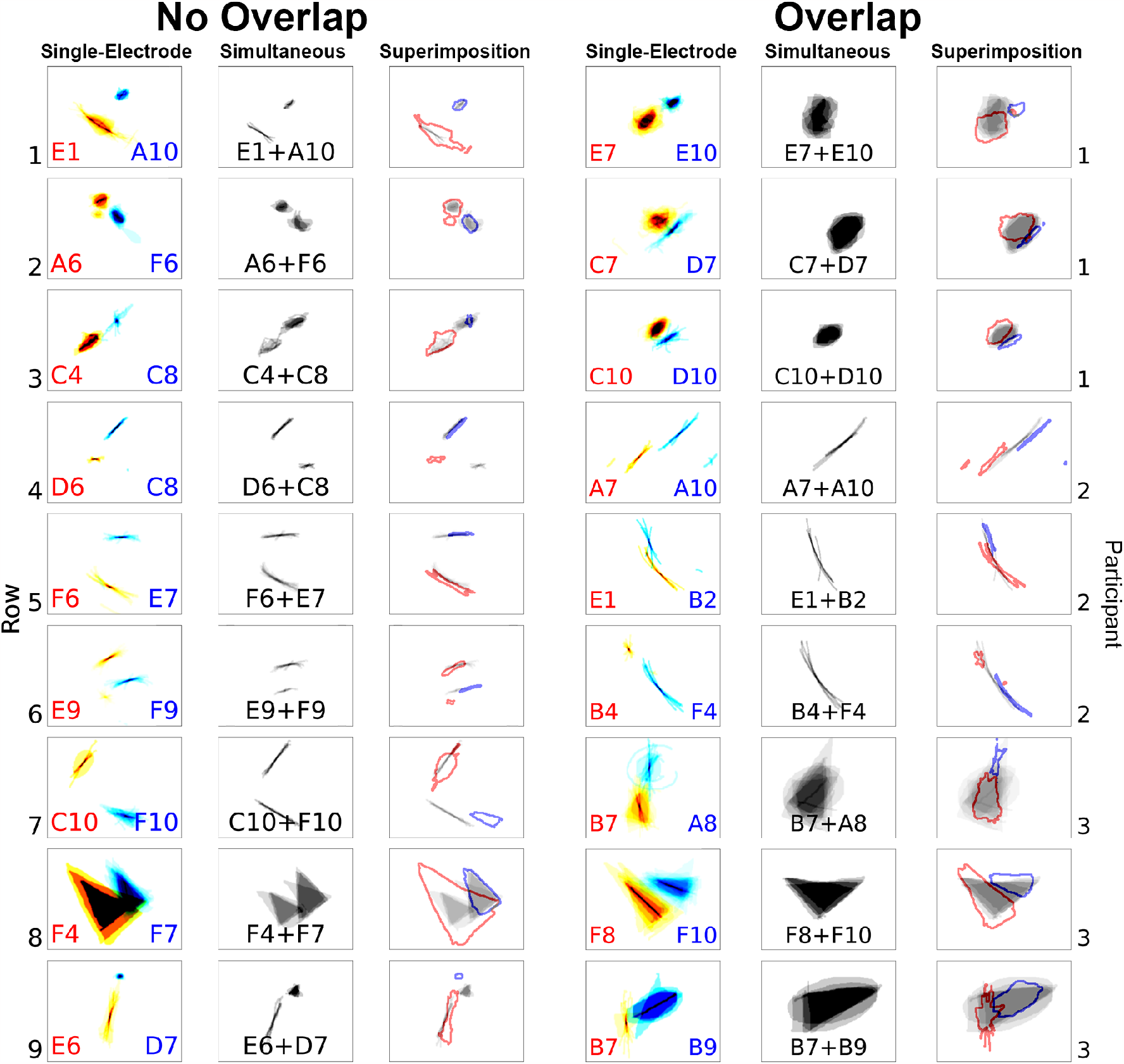
Overlaying outlines of single-electrode stimulated phosphenes onto paired-electrode stimulated phosphenes. The “No Overlap” and “Overlap” panels are organized as follows: First column: mapping two single-electrode stimulated phosphenes onto the same plot where the distance between two electrodes’ phosphenes is the scaled electrode-electrode distance. The phosphene is color-coded to match the text color of the stimulating electrode. Second column: the corresponding paired-electrode stimulated phosphenes. Third column: superimposing the outlines of phosphenes from the first column onto the phosphenes from the second column.

## References

Ahuja, A. K., Yeoh, J., Dorn, J. D., Caspi, A., Wuyyuru, V., McMahon, M. J., Humayun, M. S., Greenberg, R. J., and Dacruz, L. (2013). Factors Affecting Perceptual Threshold in Argus II Retinal Prosthesis Subjects. Transl Vis Sci Technol, 2(4):1.

Barry, M. P., Armenta Salas, M., Patel, U., Wuyyuru, V., Niketeghad, S., Bosking, W. H., Yoshor, D., Dorn, J. D., and Pouratian, N. (2020). Video-mode percepts are smaller than sums of single-electrode phosphenes with the Orion® visual cortical prosthesis. Investigative Ophthalmology & Visual Science, 61(7):927.

Beauchamp, M. S., Oswalt, D., Sun, P., Foster, B. L., Magnotti, J. F., Niketeghad, S., Pouratian, N., Bosking, W. H., and Yoshor, D. (2020). Dynamic Stimulation of Visual Cortex Produces Form Vision in Sighted and Blind Humans. Cell, 181(4):774–783.e5.

Benkrid, K., Crookes, D., and Benkrid, A. (2000). Design and FPGA implementation of a perimeter estimator. In Proceedings of the Irish Machine Vision and Image Processing Conference, pages 51–57.

Beyeler, M. (2019). Commentary: Detailed Visual Cortical Responses Generated by Retinal Sheet Transplants in Rats With Severe Retinal Degeneration. Frontiers in Neuroscience, 13.

Beyeler, M., Boynton, G., Fine, I., and Rokem, A. (2017a). pulse2percept: A Python-based simulation framework for bionic vision. In Proceedings of the 16th Python in Science Conference, pages 81–88, Austin, Texas. SciPy.

Beyeler, M., Boynton, G. M., Fine, I., and Rokem, A. (2019a). Model-Based Recommendations for Optimal Surgical Placement of Epiretinal Implants. In Shen, D., Liu, T., Peters, T. M., Staib, L. H., Essert, C., Zhou, S., Yap, P.-T., and Khan, A., editors, Medical Image Computing and Computer Assisted Intervention – MICCAI 2019, Lecture Notes in Computer Science, pages 394–402. Springer International Publishing.

Beyeler, M., Nanduri, D., Weiland, J. D., Rokem, A., Boynton, G. M., and Fine, I. (2019b). A model of ganglion axon pathways accounts for percepts elicited by retinal implants. Scientific Reports, 9(1):1–16.

Beyeler, M., Rokem, A., Boynton, G. M., and Fine, I. (2017b). Learning to see again: biological constraints on cortical plasticity and the implications for sight restoration technologies. J Neural Eng, 14(5):051003.

Bruce, A. and Beyeler, M. (2022). Greedy Optimization of Electrode Arrangement for Epiretinal Prostheses. In Medical Image Computing and Computer Assisted Intervention – MICCAI 2022: 25th International Conference, Singapore, September 18–22, 2022, Proceedings, Part VII, pages 594–603, Berlin, Heidelberg. Springer-Verlag.

Chen, S. C., Suaning, G. J., Morley, J. W., and Lovell, N. H. (2009). Simulating prosthetic vision: I. Visual models of phosphenes. Vision Research, 49(12):1493–1506.

Chen, X., Wang, F., Fernandez, E., and Roelfsema, P. R. (2020). Shape perception via a high-channel-count neuroprosthesis in monkey visual cortex. Science, 370(6521):1191–1196. Publisher: American Association for the Advancement of Science Section: Research Article.

Christie, B., Sadeghi, R., Kartha, A., Caspi, A., Tenore, F. V., Klatzky, R. L., Dagnelie, G., and Billings, S. (2022). Sequential epiretinal stimulation improves discrimination in simple shape discrimination tasks only. Journal of Neural Engineering, 19(3):036033. Publisher: IOP Publishing.

Curcio, C. A. and Allen, K. A. (1990). Topography of ganglion cells in human retina. J Comp Neurol, 300(1):5–25.

da Cruz, L., Fynes, K., Georgiadis, O., Kerby, J., Luo, Y. H., Ahmado, A., Vernon, A., Daniels, J. T., Nommiste, B., Hasan, S. M., Gooljar, S. B., Carr, A.-J. F., Vugler, A., Ramsden, C. M., Bictash, M., Fenster, M., Steer, J., Harbinson, T., Wilbrey, A., Tufail, A., Feng, G., Whitlock, M., Robson, A. G., Holder, G. E., Sagoo, M. S., Loudon, P. T., Whiting, P., and Coffey, P. J. (2018). Phase 1 clinical study of an embryonic stem cell–derived retinal pigment epithelium patch in age-related macular degeneration. Nature Biotechnology, 36(4):328–337.

de Balthasar, C., Patel, S., Roy, A., Freda, R., Greenwald, S., Horsager, A., Mahadevappa, M., Yanai, D., McMahon, M. J., Humayun, M. S., Greenberg, R. J., Weiland, J. D., and Fine, I. (2008). Factors Affecting Perceptual Thresholds in Epiretinal Prostheses. Investigative ophthalmology & visual science, 49(6):2303–2314.

de Ruyter van Steveninck, J., Güçlü, U., van Wezel, R., and van Gerven, M. (2022). End-to-end optimization of prosthetic vision. Journal of Vision, 22(2):20.

Fernández, E., Alfaro, A., Soto-Sánchez, C., Gonzalez-Lopez, P., Lozano, A. M., Peña, S., Grima, M. D., Rodil, A., Gómez, B., Chen, X., Roelfsema, P. R., Rolston, J. D., Davis, T. S., and Normann, R. A. (2021). Visual percepts evoked with an intracortical 96-channel microelectrode array inserted in human occipital cortex. The Journal of Clinical Investigation, 131(23). Publisher: American Society for Clinical Investigation.

Foik, A. T., Lean, G. A., Scholl, L. R., McLelland, B. T., Mathur, A., Aramant, R. B., Seiler, M. J., and Lyon, D. C. (2018). Detailed Visual Cortical Responses Generated by Retinal Sheet Transplants in Rats with Severe Retinal Degeneration. Journal of Neuroscience, 38(50):10709–10724. Publisher: Society for Neuroscience Section: Research Articles.

Freeman, J. and Simoncelli, E. P. (2011). Metamers of the ventral stream. Nat Neurosci, 14(9):1195–1201.

Gasparini, S. J., Llonch, S., Borsch, O., and Ader, M. (2019). Transplantation of photoreceptors into the degenerative retina: Current state and future perspectives. Progress in Retinal and Eye Research, 69:1–37.

Granley, J. and Beyeler, M. (2021). A Computational Model of Phosphene Appearance for Epiretinal Prostheses. In 2021 43rd Annual International Conference of the IEEE Engineering in Medicine Biology Society (EMBC), pages 4477–4481. ISSN: 2694-0604.

Granley, J., Relic, L., and Beyeler, M. (2022). Hybrid Neural Autoencoders for Stimulus Encoding in Visual and Other Sensory Neuroprostheses. In Advances in Neural Information Processing Systems, volume 35, pages 22671–22685.

Hamel, C. (2006). Retinitis pigmentosa. Orphanet Journal of Rare Diseases, 1(1):40.

Horsager, A., Boynton, G. M., Greenberg, R. J., and Fine, I. (2011). Temporal interactions during paired-electrode stimulation in two retinal prosthesis subjects. Invest Ophthalmol Vis Sci, 52(1):549–57.

Horsager, A., Greenwald, S. H., Weiland, J. D., Humayun, M. S., Greenberg, R. J., McMahon, M. J., Boynton, G. M., and Fine, I. (2009). Predicting Visual Sensitivity in Retinal Prosthesis Patients. Investigative Ophthalmology & Visual Science, 50(4):1483–1491.

Hou, Y., Nanduri, D., Granley, J., Weiland, J. D., and Beyeler, M. (2023). Phosphene shape elicited by paired-electrode stimulation is well predicted by single-electrode parameters for three Argus II users. Investigative Ophthalmology & Visual Science, 64(8):4613.

Hu, M.-K. (1962). Visual pattern recognition by moment invariants. IRE Transactions on Information Theory, 8(2).

Jansonius, N. M., Nevalainen, J., Selig, B., Zangwill, L. M., Sample, P. A., Budde, W. M., Jonas, J. B., Lagrèze, W. A., Airaksinen, P. J., Vonthein, R., Levin, L. A., Paetzold, J., and Schiefer, U. (2009). A mathematical description of nerve fiber bundle trajectories and their variability in the human retina. Vision Research, 49(17):2157–2163.

Luo, Y. H.-L. and da Cruz, L. (2016). The Argus® II Retinal Prosthesis System. Progress in Retinal and Eye Research, 50:89–107.

Luo, Y. H.-L., Zhong, J. J., Clemo, M., and da Cruz, L. (2016). Long-term Repeatability and Reproducibility of Phosphene Characteristics in Chronically Implanted Argus II Retinal Prosthesis Subjects. American Journal of Ophthalmology, 170:100–109.

Mahadevappa, M., Weiland, J., Yanai, D., Fine, I., Greenberg, R., and Humayun, M. (2005). Perceptual thresholds and electrode impedance in three retinal prosthesis subjects. IEEE Transactions on Neural Systems and Rehabilitation Engineering, 13(2):201–206.

McGregor, J. E. (2019). Restoring vision at the fovea. Current Opinion in Behavioral Sciences, 30:210–216.

Nanduri, D. (2011). Prosthetic vision in blind human patients: Predicting the percepts of epiretinal stimulation. PhD thesis, University of Southern California, Los Angeles, CA.

Nanduri, D., Fine, I., Horsager, A., Boynton, G. M., Humayun, M. S., Greenberg, R. J., and Weiland, J. D. (2012). Frequency and Amplitude Modulation Have Different Effects on the Percepts Elicited by Retinal Stimulation. Investigative Ophthalmology & Visual Science, 53(1):205–214.

Nanduri, D., Humayun, M. S., Greenberg, R. J., McMahon, M. J., and Weiland, J. D. (2008). Retinal prosthesis phosphene shape analysis. Conference proceedings: … Annual International Conference of the IEEE Engineering in Medicine and Biology Society. IEEE Engineering in Medicine and Biology Society. Annual Conference, 2008:1785–1788.

Oswalt, D., Bosking, W., Sun, P., Sheth, S. A., Niketeghad, S., Salas, M. A., Patel, U., Greenberg, R., Dorn, J., Pouratian, N., Beauchamp, M., and Yoshor, D. (2021). Multi-electrode stimulation evokes consistent spatial patterns of phosphenes and improves phosphene mapping in blind subjects. Brain Stimulation: Basic, Translational, and Clinical Research in Neuromodulation, 14(5):1356–1372. Publisher: Elsevier.

Perez-Yus, A., Bermudez-Cameo, J., Guerrero, J. J., and Lopez-Nicolas, G. (2017). Depth and Motion Cues with Phosphene Patterns for Prosthetic Vision. In 2017 IEEE International Conference on Computer Vision Workshops (ICCVW), pages 1516–1525. ISSN: 2473-9944.

Pogoncheff, G., Hu, Z., Rokem, A., and Beyeler, M. (2023). Explainable Machine Learning Predictions of Perceptual Sensitivity for Retinal Prostheses. Pages: 2023.02.09.23285633.

Relic, L., Zhang, B., Tuan, Y.-L., and Beyeler, M. (2022). Deep Learn-ing–Based Perceptual Stimulus Encoder for Bionic Vision. In Augmented Humans 2022, AHs 2022, pages 323–325, New York, NY, USA. Association for Computing Machinery.

Rizzo, J. F., Wyatt, J., Loewenstein, J., Kelly, S., and Shire, D. (2003). Perceptual Efficacy of Electrical Stimulation of Human Retina with a Microelectrode Array during Short-Term Surgical Trials. Investigative Ophthalmology & Visual Science, 44(12):5362–5369. Publisher: The Association for Research in Vision and Ophthalmology.

Roelfsema, P. R. (2023). Solving the binding problem: Assemblies form when neurons enhance their firing rate—they don’t need to oscillate or synchronize. Neuron, 111(7):1003–1019. Publisher: Elsevier.

Russell, S., Bennett, J., Wellman, J. A., Chung, D. C., Yu, Z.-F., Tillman, A., Wittes, J., Pappas, J., Elci, O., McCague, S., Cross, D., Marshall, K. A., Walshire, J., Kehoe, T. L., Reichert, H., Davis, M., Raffini, L., George, L. A., Hudson, F. P., Dingfield, L., Zhu, X., Haller, J. A., Sohn, E. H., Mahajan, V. B., Pfeifer, W., Weckmann, M., Johnson, C., Gewaily, D., Drack, A., Stone, E., Wachtel, K., Simonelli, F., Leroy, B. P., Wright, J. F., High, K. A., and Maguire, A. M. (2017). Efficacy and safety of voretigene neparvovec (AAV2-hRPE65v2) in patients with RPE65-mediated inherited retinal dystrophy: a randomised, controlled, open-label, phase 3 trial. The Lancet, 390(10097):849–860. Publisher: Elsevier.

Sanchez-Garcia, M., Martinez-Cantin, R., and Guerrero, J. (2019). Indoor Scenes Understanding for Visual Prosthesis with Fully Convolutional Networks:. In Proceedings of the 14th International Joint Conference on Computer Vision, Imaging and Computer Graphics Theory and Applications, pages 218–225, Prague, Czech Republic. SCITEPRESS - Science and Technology Publications.

Shivdasani, M. N., Sinclair, N. C., Gillespie, L. N., Petoe, M. A., Titchener, S. A., Fallon, J. B., Perera, T., Pardinas-Diaz, D., Barnes, N. M., Blamey, P. J., and for the Bionic Vision Australia Consortium (2017). Identification of Characters and Localization of Images Using Direct Multiple-Electrode Stimulation With a Suprachoroidal Retinal Prosthesis. Investigative Ophthalmology & Visual Science, 58(10):3962–3974.

Sinclair, N. C., Shivdasani, M. N., Perera, T., Gillespie, L. N., McDermott, H. J., Ayton, L. N., and Blamey, P. J. (2016). The Appearance of Phosphenes Elicited Using a Suprachoroidal Retinal Prosthesis. Investigative Ophthalmology & Visual Science, 57(11):4948–4961.

Song, X., Qiu, S., Shivdasani, M. N., Zhou, F., Liu, Z., Ma, S., Chai, X., Chen, Y., Cai, X., Guo, T., and Li, L. (2022). An in-silico analysis of electrically-evoked responses of midget and parasol retinal ganglion cells in different retinal regions. Journal of Neural Engineering.

Spencer, M. J., Kameneva, T., Grayden, D. B., Meffin, H., and Burkitt, A. N. (2019). Global activity shaping strategies for a retinal implant. Journal of Neural Engineering, 16(2):026008. Publisher: IOP Publishing.

Stingl, K., Bartz-Schmidt, K. U., Besch, D., Chee, C. K., Cottriall, C. L., Gekeler, F., Groppe, M., Jackson, T. L., MacLaren, R. E., Koitschev, A., Kusnyerik, A., Neffendorf, J., Nemeth, J., Naeem, M. A. N., Peters, T., Ramsden, J. D., Sachs, H., Simpson, A., Singh, M. S., Wilhelm, B., Wong, D., and Zrenner, E. (2015). Subretinal Visual Implant Alpha IMS – Clinical trial interim report. Vision Research, 111:149–160.

Stingl, K., Bartz-Schmidt, K.-U., Gekeler, F., Kusnyerik, A., Sachs, H., and Zrenner, E. (2013). Functional Outcome in Subretinal Electronic Implants Depends on Foveal Eccentricity. Investigative Ophthalmology & Visual Science, 54(12):7658–7665.

Weiland, J. D., Walston, S. T., and Humayun, M. S. (2016). Electrical Stimulation of the Retina to Produce Artificial Vision. Annual Review of Vision Science, 2(1):273–294.

Wilke, R., Gabel, V.-P., Sachs, H., Bartz Schmidt, K.-U., Gekeler, F., Besch, D., Szurman, P., Stett, A., Wilhelm, B., Peters, T., Harscher, A., Greppmaier, U., Kibbel, S., Benav, H., Bruckmann, A., Stingl, K., Kusnyerik, A., and Zrenner, E. (2011a). Spatial resolution and perception of patterns mediated by a subretinal 16-electrode array in patients blinded by hereditary retinal dystrophies. Invest Ophthalmol Vis Sci, 52(8):5995–6003.

Wilke, R. G. H., Moghadam, G. K., Lovell, N. H., Suaning, G. J., and Dokos, S. (2011b). Electric crosstalk impairs spatial resolution of multi-electrode arrays in retinal implants. Journal of Neural Engineering, 8(4):046016. Publisher: IOP Publishing.

Yücel, E. I., Sadeghi, R., Kartha, A., Montezuma, S. R., Dagnelie, G., Rokem, A., Boynton, G. M., Fine, I., and Beyeler, M. (2022). Factors affecting two-point discrimination in Argus II patients. Frontiers in Neuroscience, 16.

Zrenner, E., Bartz-Schmidt, K. U., Benav, H., Besch, D., Bruckmann, A., Gabel, V.-P., Gekeler, F., Greppmaier, U., Harscher, A., Kibbel, S., Koch, J., Kusnyerik, A., Peters, T., Stingl, K., Sachs, H., Stett, A., Szurman, P., Wilhelm, B., and Wilke, R. (2010). Subretinal electronic chips allow blind patients to read letters and combine them to words. Proceedings of the Royal Society B: Biological Sciences, 278(1711):1489–1497. Publisher: Royal Society.

